# An accurate hierarchical model to forecast diverse seasonal infectious diseases

**DOI:** 10.1101/2025.03.03.25323259

**Authors:** B. K. M. Case, Mariah Victoria Salcedo, Spencer J. Fox

## Abstract

Since 2021, the seasonal tripledemic composed of COVID-19, influenza, and respiratory syncytial virus (RSV) has threatened healthcare capacity globally. Short-term forecasts can provide public health officials and healthcare leaders time to effectively respond to epidemics, but many forecast approaches are bespoke to specific diseases or localities. We present a hierarchical forecast model that flexibly accounts for spatial and seasonal transmission dynamics and test its performance on hospital admissions in the United States over two years. The model outcompetes a baseline forecast model by 42%, 44%, and 41% for COVID-19, influenza, and RSV respectively, and it was the top individual forecast model in the 2023-2024 CDC FluSight forecast challenge. We use it to quantify the single-peaked timing and shape for influenza and RSV epidemics and the biannual seasonality of COVID-19. Additionally, we estimate regional disease burden differences across the country with higher burden in the South and lower burden in the West and Northeast. Given its flexible nature and robust performance, our model provides a straightforward way to expand forecasting to additional regions and for other seasonal diseases such as Dengue virus or malaria.

## Introduction

Prior to the emergence of novel SARS-CoV-2 coronavirus (COVID-19) in early 2020, the respiratory virus season in the United States was composed of respiratory syncytial virus (RSV) and influenza, which combined to cause 5,400-30,800 annual deaths on average [1,2] and periodically overwhelmed healthcare systems such as in 2017-2018 during the H3N2 influenza epidemic [3–5]. Beginning with the 2021 to 2022 respiratory virus season COVID-19, RSV, and influenza have co-circulated and now cause a combined average of approximately 4 million hospitalizations in the United States [6–11], with the annual potential to overwhelm healthcare systems with the emergence of a more infectious or severe variant or with a mismatch between the selected vaccine and circulating strains [12].

Short-term outbreak forecasts, predictions of epidemiological burden one to four weeks in advance, became a critical public health tool during the COVID-19 pandemic and are used to inform more timely interventions and the allocation of critical resources, particularly when resources are strained during an epidemic surge [13]. Collaborative forecast hubs are the current standard for real-time forecast efforts, because they provide a way to coordinate the production, submission, evaluation, and communication of forecasts and allow for the production of aggregated or *ensemble* forecasts that tend to outperform individual forecast models [14–16]. The Centers for Disease Control and Prevention (CDC) coordinated such efforts for regional influenza forecasts annually beginning in 2013 as part of the FluSight challenge [17–19], and as of 2024, similar efforts exist for forecasting COVID-19 and RSV both domestically and internationally [17,20–22]. Although forecasting efforts have expanded rapidly over the past decade and demonstrated clear public health utility, their performance remains inconsistent, highlighting the need for further improvement [17,23].

Successful forecast models have historically been developed and optimized for a specific disease, data set, or spatial context. For example, forecast models may be built to mechanistically capture humidity- or evolutionary- driven transmission dynamics [24–27], to derive statistical relationships between large digital data streams like mobility or electronic health records and the epidemiological target of interest [28,29], or to leverage long-running historical time-series for training accurate forecast models [30–33]. Such approaches, though, often require expert modification and fine-tuning to apply to alternative diseases, time periods, or locations, a process that may be impossible for data-poor settings such as during outbreaks of emergent or neglected diseases.

Here we developed a flexible Bayesian spatiotemporal hierarchical model to address the challenges presented by limited training data and bespoke model configuration for forecasting seasonal infectious diseases. Our model leverages salient features of seasonal epidemics including that they tend to occur at the same time each year, that specific seasons are worse than others, and that nearby regions tend to have similar dynamics to one another [34–36]. To demonstrate the flexibility of the model, we applied it to forecast three common seasonal respiratory diseases with distinct epidemiological dynamics: (1) RSV that has winter seasonality and an estimated 97% of hospitalizations occurring in children under five [37], (2) influenza that has winter seasonality and the highest hospitalization rates in adults older than 65 [38,39], and (3) COVID-19 that primarily hospitalizes adults older than 65 and has significant spatial heterogeneity and multiple epidemic waves a year [40–42]. Our results show that the model has robust forecast performance for all three diseases and that it outperforms top alternative models in real-time head-to-head comparisons. Additionally, the inferred model parameters provide quantitative insights into the co-circulation of the three viruses in the US.

## Results

To motivate the design of our seasonal disease forecast model, we first compare the spatiotemporal dynamics of influenza, COVID-19, and RSV hospitalizations (Figure 1). The incidence rates for all three diseases show stark seasonal patterns (Figure 1A-1C), with major epidemic peaks for all occurring between mid-December and late-February (corresponding to respiratory virus season week 10 and 25 respectively), and an additional COVID-19 peak in the late summer (Figure 1A-C). When we remove the seasonal trend (*i.e.* the average weekly hospitalization rate across all states and years) from the state-specific time-series, significant variability remains (Figure 1D-F). For influenza and RSV for example, the 2022-2023 season hospitalization rates were twice the seasonal average for many states, suggesting an abnormally large seasonal epidemic. Abnormally high rates early followed by abnormally low rates later highlight the early wave of RSV that occurred that winter. Geographic proximity impacts the remaining variability in trends across states, with the hospitalization rates of nearby states tending to be more correlated with one another than distant ones (Figure 1G-I). The pattern is especially stark for COVID-19, where states sharing a border have a median correlation of 85% (95% CI: 82%-86%) compared to just 37% (95% CI: 25%-43%) for those eight states apart (Figure 1G).

**Figure 1:**
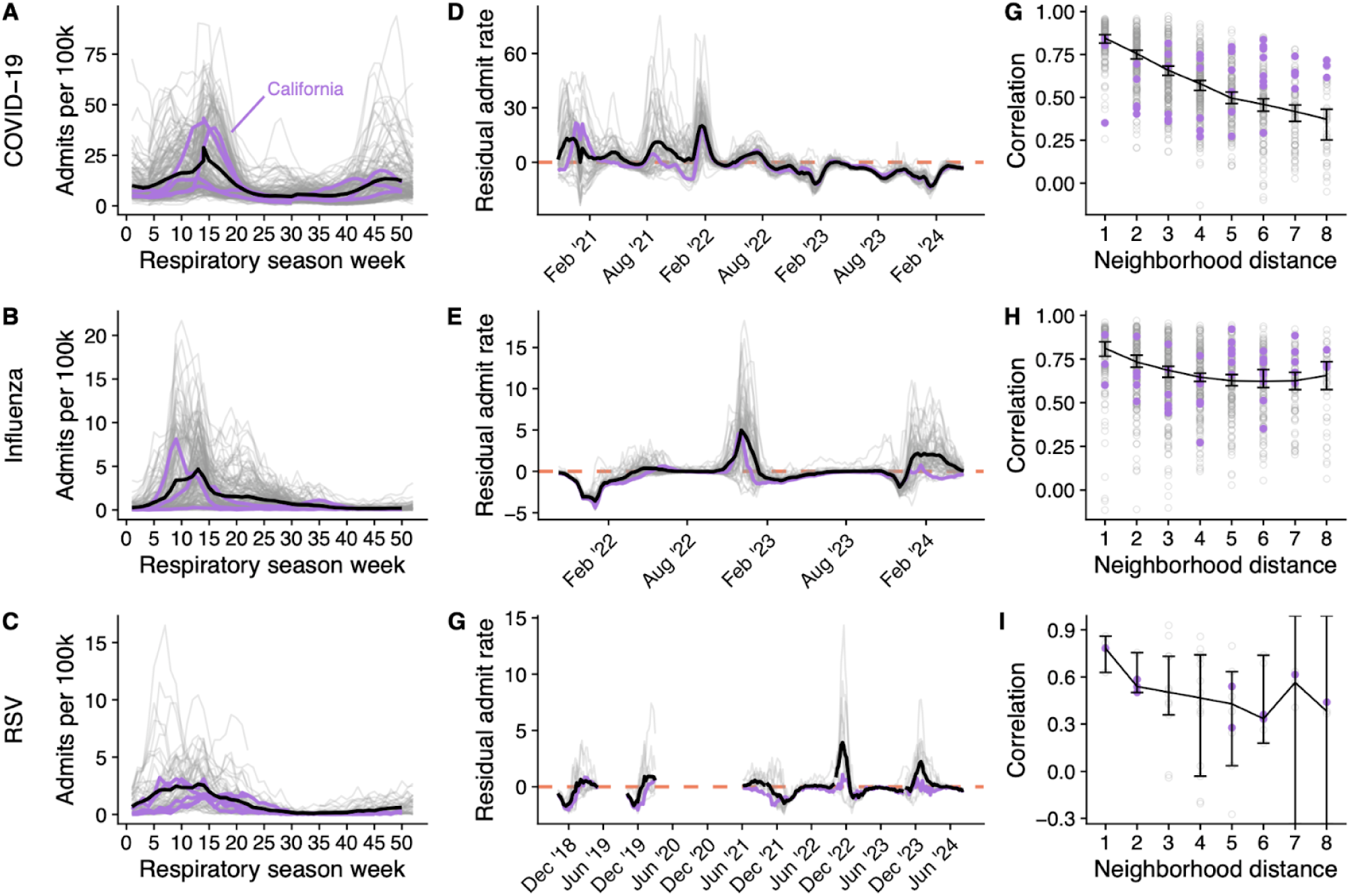
Seasonal, temporal, and spatial epidemiological dynamics for COVID-19, influenza, and RSV hospital admissions at the state-level. **(A-C)**: Hospitalization rates per 100,000 individuals for each state, season, and respiratory virus (grey lines) by the week of the respiratory virus season where zero corresponds to epidemiological week 40. The black line indicates the average, illustrating the seasonal trend. **(D-F)** Residual hospitalization rates for each date and state (grey lines) alongside the average across all states (black line). For each date and state, we subtract the seasonal trend from that week to create the residual rate. The dashed line indicates average seasonal patterns (Y=0) with positive values indicating higher than average burden and negative values indicating lower than average burden for that week. **(G-I)** Pairwise state Pearson correlation for hospital admission rates by the shortest-path distance between state boundaries (grey points) alongside the median and 95% confidence interval on the median (black line and error bars respectively). For all panels, data for California are highlighted in purple. COVID-19 and influenza data are obtained from the CDC’s NHSN dataset, while the RSV data are from CDC’s RSV-NET [10,44]. Note that we removed data for RSV and influenza between early 2020 and mid 2021 to account for abnormally small seasons during COVID-19 and that RSV-NET only provided estimates from epidemiological weeks 40 through 18 until 2021.

Motivated by these observations, we developed a Bayesian hierarchical spatiotemporal model that decomposes state hospital admissions into a seasonal effect that is shared across states and a short-term effect that captures non-seasonal, week-to-week variation. The short-term effect is made up of a main effect that accounts for non-seasonal patterns common to all states and an interaction effect that accounts for spatiotemporal correlations. We tested the real-time forecast performance of the model, named INFLAenza, by submitting probabilistic predictions for observed influenza hospitalizations 1-4 weeks ahead (corresponding to horizons 0-3) for all 50 states, Puerto Rico, District of Columbia, and nationally to the 2023-2024 FluSight forecasting hub from October 14, 2023 to May 4, 2024. We summarize the forecast performance using the two standard metrics of collaborative forecast hubs: (1) the weighted interval score (WIS) that is the primary performance metric and summarizes both the accuracy and precision of probabilistic forecasts and (2) the prediction interval coverage (PIC) that measures the proportion of forecasts that accurately capture the underlying data for a given interval width (e.g. a 95% prediction interval) [18,43]. Rather than use the raw WIS score, we estimate the relative (rWIS) performance against a baseline forecast model that is a random walk of order one and serves as a forecast performance reference point, with lower rWIS values indicating better performance and values below one indicating that the model outperformed the baseline model.

Out of the 24 eligible models who submitted at least 80% of all forecasts, INFLAenza was the 4th best overall and the top individual forecast model by rWIS with a score of 0.76, indicating a 24% improvement compared to the FluSight baseline model (Table 1). INFLAenza was 7.3% better than the next best individual forecast model and it was only 4.1% worse than the FluSight ensemble model, which has historically been the top performing model. INFLAenza achieved PIC values of 47.9% and 90.8% compared to the nominal 50% and 95% expectations respectively and was in line with other top forecast models.

**Table 1:**
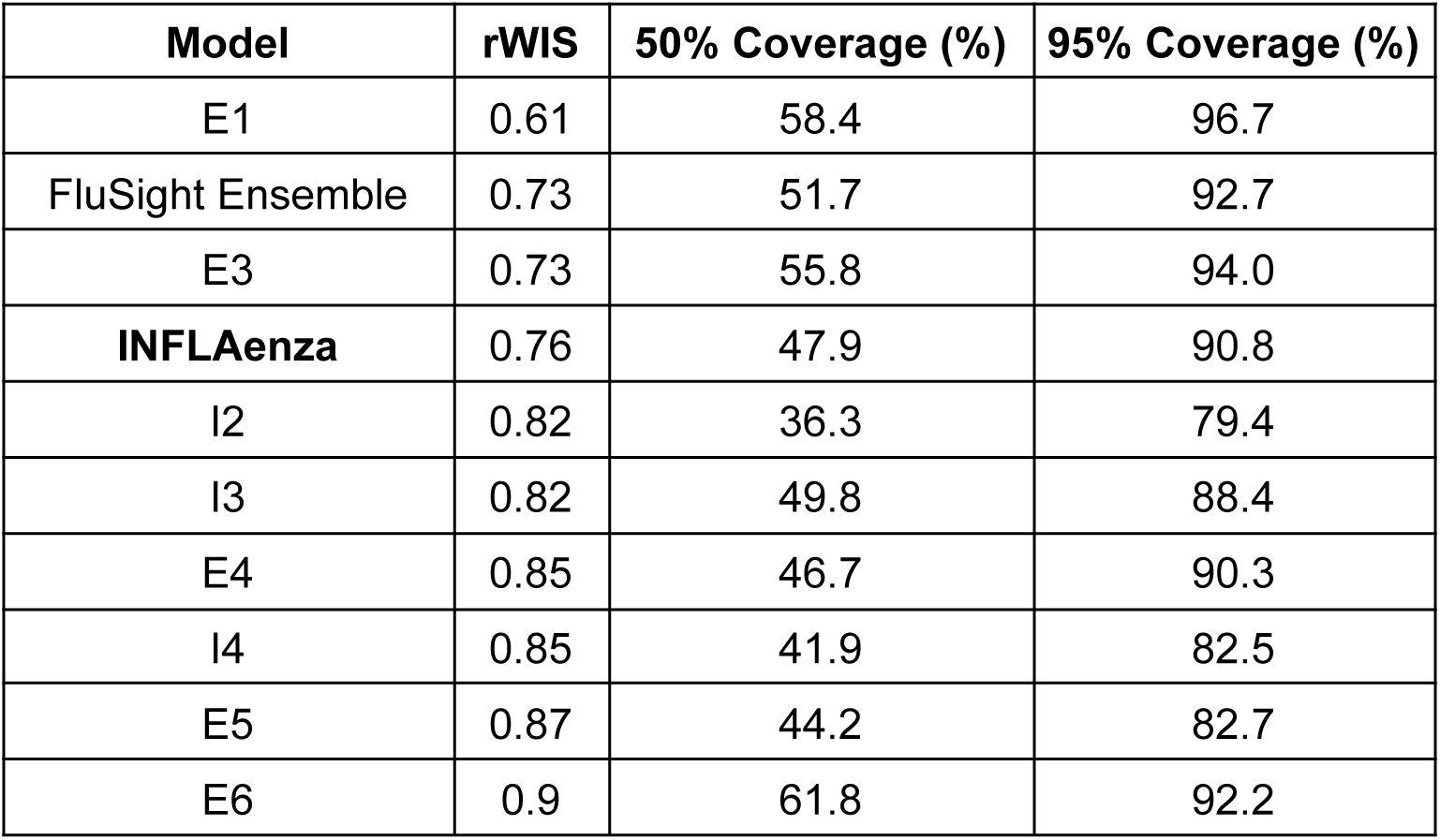
Performance metrics of the top ten models in the 2023-24 FluSight challenge. Models are ranked according to their average relative Weighted Interval Score (rWIS) compared with a reference baseline model across all forecasted dates, regions, and horizons. We anonymize each of the models other than INFLAenza and the FluSight Ensemble with a letter and number, where ensemble and individual models are identified with an “E” or “I” respectively.

Comparing the relative performance of the model through time, we find that INFLAenza outperformed or performed within 5% of the FluSight baseline model for all weeks except for during the peak on December 30, 2023 and the first sign of a shoulder to the season on January 27, 2024 (Figure 2A). Our performance against the FluSight ensemble model was more mixed, with INFLAenza outperforming it by 15.9% during the first six weeks of the season and underperforming it by 12.5% during the rest of the season. INFLAenza performed similarly when viewed at the state-level, matching or outperforming the baseline forecast model for 90.4% of states, while matching or outperforming the ensemble model for 50% of all states (Figure 2B).

**Figure 2:**
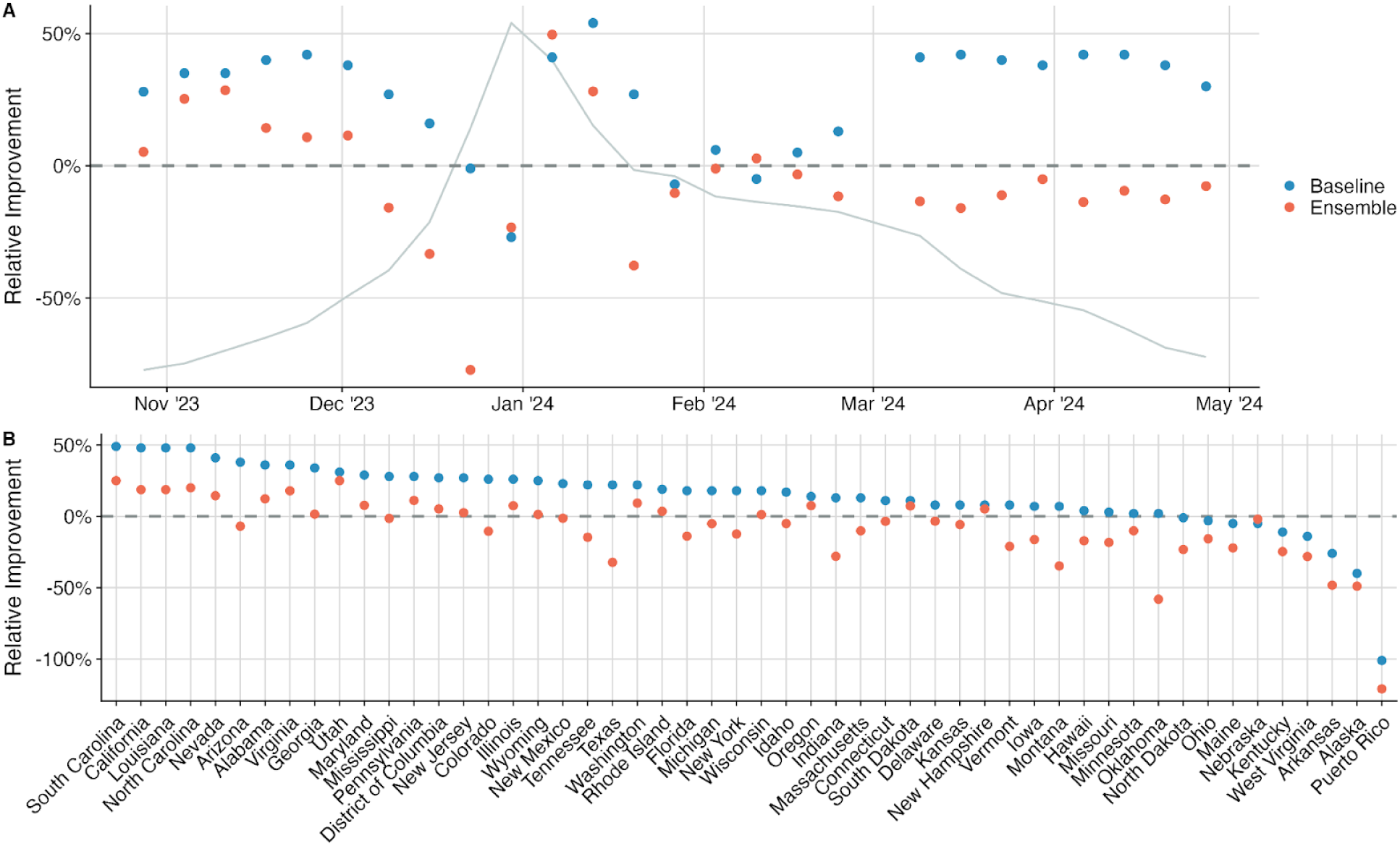
Temporal and regional forecast performance of the INFLAenza model compared against the baseline and FluSight ensemble models during the 2023-2024 influenza season. **(A)** Relative percent improvement of the INFLAenza model rWIS compared to that of the FluSight baseline (blue points) or the FluSight ensemble (orange points) across all 50 states, Puerto Rico, and District of Columbia and the 1-4 week(s) ahead predictions for each forecast date. Timing of the performance is compared against the timing of the national influenza hospital admission epidemiological curve (grey line). **(B)** Relative percent improvement of the INFLAenza model compared to the FluSight baseline (blue points) or the FluSight ensemble (orange points) across the full time period and the 1-4 week(s) ahead predictions for each forecasted region. For both panels positive values indicate that INFLAenza outperformed the compared model.

INFLAenza consistently beat the FluSight baseline model across all forecast horizons, though it performed below the ensemble model by 1.3%, 2.7%, 4.2%, and 8.5%, for the zero-, one-, two-, and three-week horizons ahead predictions respectively (Figure S1). Overall, we find INFLAenza performed similarly when measured by PIC, outperforming the baseline model and nearly matching or just underperforming against the FluSight ensemble (Figure S2-S4).

To test the performance of INFLAenza across multiple diseases and seasons we produced retrospective forecasts of hospital admissions for COVID-19, influenza, and RSV independently but with the same underlying model. Specifically, we followed their respective hub forecasting formats and produced 1-4 week ahead quantile forecasts every week from September 09, 2022 to April 27, 2024 for all 50 states, the District of Columbia, and Puerto Rico for COVID-19 and influenza, and for the twelve states available from RSV-net (CA, CO, CT, GA, MD, MI, MN, NM, NY, OR, TN, and UT) for RSV [18,20,22]. Overall, INFLAenza’s forecasts performed 41.7%, 44%, and 40.6% better compared to the baseline model for COVID-19, influenza, and RSV respectively, with model forecasts aligning well with the underlying epidemiological trends (Figure 3A-C). We find that forecasts were well calibrated for each of the diseases, with 95% PIC estimates for COVID-19, influenza, and RSV of 94.5%, 88.5%, and 96.7%, respectively. INFLAenza performs 10.8% and 7.9% better compared to reduced models that lacked either the seasonal component or spatial component (*i.e.* fitting the model independently to each state) respectively, on average across the three diseases (Table S1).

**Figure 3:**
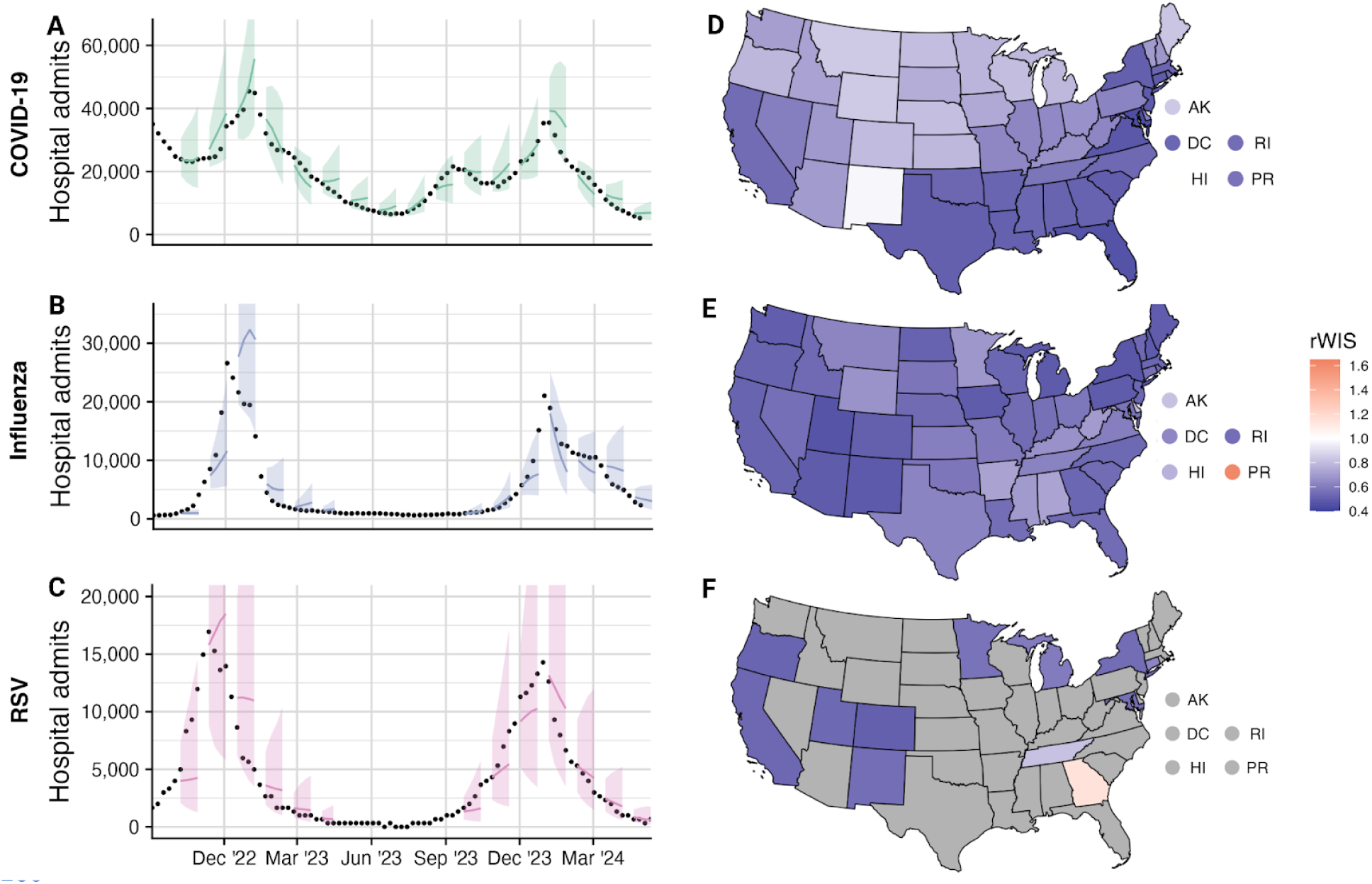
Retrospective forecast performance of the INFLAenza model for COVID-19, influenza, and RSV from September 09, 2022 to April 27, 2024. **(A-C)** National-level 1-4 week ahead mean forecasts and 95% prediction intervals for COVID-19, influenza, and RSV (lines and ribbons) compared against the observed NHSN and RSV-Net hospital admission observations (black points) [10,44]. **(D-F)** Forecast performance for all 50 states, the District of Columbia, and Puerto Rico relative to the reference baseline model (rWIS). Lower values indicate improved performance with values below 1 indicating better performance than the baseline model. Regions where data are unavailable are colored grey.

In comparing performance of the forecast model by state, INFLAenza outperformed the baseline model for all diseases and regions except for RSV in Georgia and influenza in Puerto Rico (Figure 3D-F). INFLAenza performance is low for these regions due to their distinct underlying seasonal dynamics compared to the rest of the country (Figure S5). For COVID-19, we find distinct spatial patterns in INFLAenza performance, with better performance in the Southeast and Northeast compared to the West and Midwest (Figure 3D). Comparing performance between US Census regions over time, we find these patterns are driven by differences in relative performance during the 2022/23 respiratory season (Figure S6). During this time, states in the Midwest and West experienced an earlier winter peak in COVID-19 hospitalizations, followed by a prolonged spring shoulder, while states in the South closely followed the national trend (Figure S6). The 95% PIC is similar across regions to the overall performance with values of 94.5% (95% CI: 83.8-100%), 88.5% (95% CI: 79.5-99.2%), 96.7% (95% CI: 93.1-99.3%) for COVID-19, influenza, and RSV respectively (Figure S7). We find that rWIS and PIC performance by date and horizon for all three diseases matches the real-time influenza results, with strong performance compared to the baseline for all forecast horizons and all dates apart from a handful of weeks during epidemic peaks (Figure S8-S15).

We estimate and compare the disease-specific spatiotemporal dynamics of COVID-19, influenza, and RSV using the fitted INFLAenza forecast models using all data until April 27, 2024. The posterior distribution for the seasonal effect indicates the overall trend in hospital admissions over each year as a function of the week of the respiratory virus season (Figure 4A). Our estimates demonstrate that all three diseases experience their largest burden in the winter, though they differ in their overall shapes: influenza and RSV both have sharp winter peaks, with influenza having a late winter shoulder that elongates risks, while COVID-19 seasonality is flatter overall but bimodal, with an additional smaller peak in the late summer. Overall, we find that COVID-19 rates increase first, followed by RSV, followed by influenza with estimated timing that the seasonality increases significantly above zero in epidemiological week 33 (95 % Cr: 31-44), 41 (95% CrI: 40-42), and 45 (95% CrI: 43-46), respectively. We also estimate similar peak timing across diseases, though RSV and influenza peak slightly before COVID-19, with peak epidemiological weeks estimated to be 52 (95% CrI: 49-1), 52 (95% CrI: 51-52), and 1 (95% CrI: 52-2) respectively. The posterior distributions of the short-term main effect indicate dates that depart from the seasonal trend common to all states (Figure 4B). Influenza has the most distinct pattern between seasons: for the 2021-2022 season, rates began lower than the seasonal trend during the Omicron epidemic and ended above the seasonal trend; for the 2022-2023 season, rates had the opposite pattern with initially higher rates than the seasonal trend followed by a rapid drop; and the 2023-2024 season was entirely above the seasonal trend. For COVID-19, our model suggests that hospital admission rates have gradually declined following the late 2021 Omicron epidemic. Finally, for RSV, the values are near zero with little variability, indicating non-seasonal, short-term variation in hospitalizations for RSV is better explained at the individual state level than a common trend shared between all locations.

**Figure 4:**
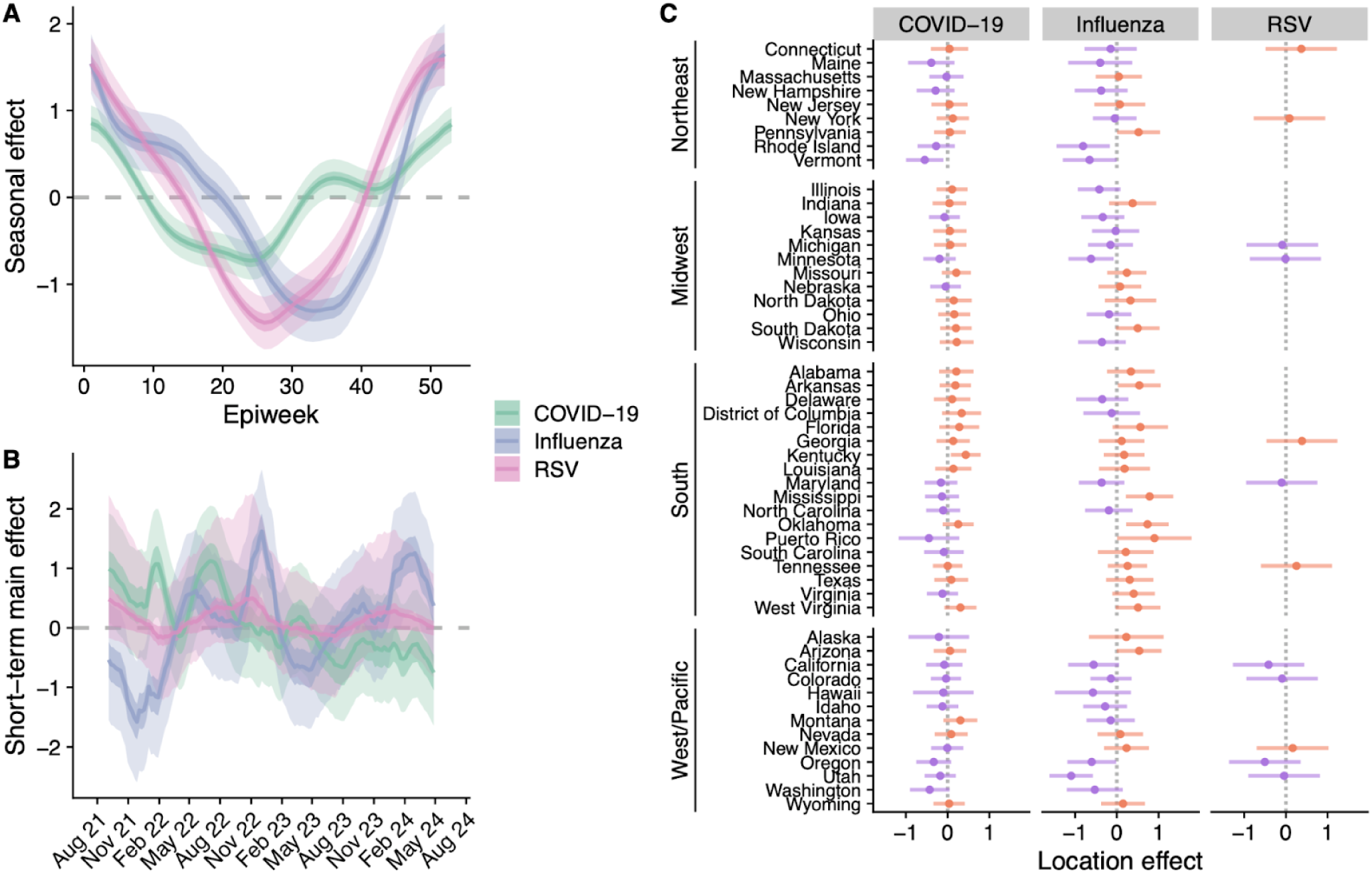
INFLAenza posterior estimates for disease-specific spatiotemporal dynamics. **(A)** Posterior mean seasonal effect (colored lines, *φ* in Eq. (2)) with 50% and 95% credible intervals (colored ribbons) as a function of epidemiological week. Values below zero indicate hospital admission rates tended to be lower for that week across states and years. **(B)** Posterior mean of the temporal main effect (colored lines, *α*) alongside the 50% and 95% credible intervals (colored ribbons). Values below zero indicate hospital rates tended to be lower than the seasonal average across all states for that date. Values for RSV and COVID-19 prior to the training period of influenza are not shown. **(C)** Posterior mean location-specific intercepts (points, *β* in Eq. (2)) and 95% credible intervals (horizontal lines) for each disease. Values are colored according to whether the posterior mean is above zero (orange) or below zero (purple). Values below zero indicate an overall lower rate of hospital admissions compared to the national average throughout the time series. Posterior estimates are derived using all training data up until April 27, 2024.

The posterior distributions for the location intercepts indicate state-specific deviations from the countrywide average, with negative and positive values indicating overall lower and higher hospital rates throughout the state’s time series compared to the national average respectively. In comparing the state-specific intercepts, we find that in general, influenza disease burden across states is more variable than COVID-19 or RSV, with 81% of influenza states having 95% credible intervals containing zero, compared to 96% and 100% for COVID-19 and RSV respectively (Figure 4C). For influenza, we find strong support that hospitalization rates are higher in the South and moderate support that rates are lower in the Northeast and the Pacific West, while for COVID-19, we find moderate support that rates are lower in the northeast (Figure S16). We also find a positive correlation of of 0.42, 0.61, and 0.70 for the location-specific intercepts between COVID-19 and influenza, influenza and RSV, and COVID-19 and RSV respectively, suggesting that states with a high (low) burden of one disease tend to have a high (low) burden of the other diseases (Figure S17). However, the correlations do not account for uncertainty on the location intercepts, nor the impact of short-term differences in hospitalizations captured by the spatiotemporal interaction component of the model.

## Discussion

In this paper we have developed a new, disease-agnostic Bayesian hierarchical spatiotemporal forecasting model for seasonal infectious diseases and applied it to forecasting COVID-19, influenza, and RSV hospital admissions in the United States over multiple years.

The model outperformed all other individual forecast models in real-time head-to-head comparisons during the 2023-2024 influenza season and performed almost on par with the current gold standard ensemble forecast model (Table 1) [15]. Through retrospective evaluation, we found that the model outperforms the standard baseline forecast reference model by an average of 42% across all three diseases, suggesting robust forecast performance for diseases with distinct spatiotemporal epidemiological dynamics (Figure 3). We also used the inferred model components to quantify, previously undiscussed, seasonal and spatial patterns across the three diseases that may inform future public health interventions (Figure 4). Taken together, our results suggest that the INFLAenza model can be applied nearly off-the-shelf to accurately forecast seasonal infectious diseases in multiple regions simultaneously, opening the door to rapidly expand disease forecast efforts to new locales with different seasonal patterns and additional high impact seasonal diseases such as Dengue virus or malaria.

There are a number of existing modeling and forecasting approaches that decompose the time series into seasonal and short-term trends [45–47]. While such models are flexible and simple to apply to forecasting seasonal disease dynamics, we note that common approaches incorporating seasonality with splines or autoregressive techniques, such as SARIMA or structural equation models, have had limited success in collaborative disease forecasting efforts thus far [18,23,48,49]. We therefore believe our model is unique in incorporating these dynamics and delivering robust forecast performance on multiple diseases nearly off-the-shelf. A similar approach that used seasonal and short-term random walks alongside spatial information sharing, *Dante*, was the top performing model in the 2018/19 FluSight challenge [50]. The success of both INFLAenza and Dante illustrates the strength of a hierarchical approach for capturing key spatiotemporal dependencies for both fine-scale and aggregated patterns. INFLAenza adds an additional ability to explicitly incorporate spatial correlations between nearby localities, which may be particularly important for diseases with strong spatial heterogeneity like COVID-19 (Figure 1, Table S1). Additionally, INFLAenza produces state- and national forecasts in minutes or less, whereas Dante may take hours, limiting its potential scalability [51].

While INFLAenza outperformed the baseline model by at least 20% across all of the examined forecasts, we found that it performed better in our retrospective analysis compared to the real-time forecasts (42% improvement compared to 25%). One plausible explanation is that the FluSight hub computes rWIS in a slightly different manner, using a pairwise ranking scheme between all models in the hub to account for the fact that not all teams submit forecasts for all locations and weeks [18]. An additional explanation is that real-time forecasts must contend not only with epidemiological dynamics, but also with reporting updates and lags which alter recent observations and may hurt performance relative to the baseline model [52,53]. INFLAenza’s real-time relative performance against the baseline model may therefore be improved through integration with a nowcasting component that anticipates current and future reporting delays, potentially through harnessing the NHSN provided estimates of the percentage of reporting hospitals by date [44,54–56].

We estimate a biannual nature to COVID-19 hospitalization rates in the United States with key peaks in the winter and late summer (Figure 4A), contrasting with early expectations for annual or biennial endemic COVID-19 patterns similar to other coronaviruses [57]. The summer epidemic wave in 2020 was largely attributed to policy and behavioral relaxation following strict lockdowns in the spring of 2020 [28,58], while the 2021 summer wave was attributed to the emergence of the B.1.617.2 (Delta) variant [59]. However, our estimates are consistent with the idea that biannual dynamics may continue to be a part of the COVID-19 seasonal cycle, aligning with the observed hospitalization rates since the summer of 2020 (Figure S18). While we cannot determine the cause of this observed seasonality, previous work has suggested that behavioral-driven changes during the summer months may lead to more indoor interactions and increase airborne transmission risks similarly to colder months [60]. Alternatively, rapid COVID-19 variant evolution and immune waning could combine to explain this observed biannual pattern [42,61]. Future work is necessary to disentangle these possibilities and improve our ability to anticipate long-term COVID-19 dynamics.

Comparing the seasonality curves across diseases, we find overlapping transmission peaks for COVID-19, influenza, and RSV (Figure 4A), suggesting that they may have shared seasonality drivers such as population mobility or humidity [62–64]. Unlike COVID-19, influenza and RSV show strong unimodal seasonality, with RSV epidemics starting significantly before influenza epidemics (Figure 4A). The earlier emergence of RSV may be driven by school reopenings and household transmission to highest risk children [37,65], though school transmission is known to contribute significantly to seasonal influenza transmission as well [66]. While it is too soon to tell how the long-term co-circulation dynamics of COVID-19, influenza, and RSV will progress, our estimates highlight consistent recent seasonal patterns that can be used for future seasonal predictions.

The estimated location-specific intercepts suggest that hospitalization rates for COVID-19 and influenza tend to be higher among Southern states compared to those in the Northeast and Pacific West and that there is a positive correlation across disease-specific burden rates (Figure 4C and Figures S17). These correlations across the diseases could be explained by regional biases or differences in (1) respiratory disease surveillance system reporting rates, (2) underlying population risk factors such as age or comorbidities, or (3) the availability and uptake of non-pharmaceutical or pharmaceutical interventions like antiviral drugs and vaccinations [67–71]. Improved understanding of these possible explanations may be used to inform targeted interventions across the country in future years.

Despite the strong forecast performance of the INFLAenza model over multiple years retrospectively, we were only able to test its real-time performance on a single season. Additional years of data are needed to continually validate its performance in real-time settings. There are also a number of limitations that could be addressed to further improve the model. First, the model assumes a shared seasonal term across the whole geographic region, reducing its performance in regions such as Puerto Rico that have distinct seasonal trends. Adding regional seasonality to the model for geographically distinct locations may further improve the models performance and would allow for simultaneous forecasts of regional dynamics across hemispheres. Second, we model spatial connectivity based on the shortest-path distance between state borders, through proximity between locations could alternatively be measured in different ways such as mobility patterns. Both metrics have previously been shown to be strong predictors of spatial epidemiology, but further work is necessary to test their respective forecast performance [72–75]. Third, we do not include any exogenous data streams such as climatic, behavioral, or epidemiological variables that have previously been shown to aid forecast efforts [26,76]. We avoided such additional data to keep our model as broadly applicable as possible, though our forecasts may benefit from fine-tuning the inclusion of disease-specific early warning signals such as the estimated disease evolution or the match between the vaccine and circulating variants [77–79]. Finally, given the similar transmission pathways and co-circulating nature of COVID-19, RSV, and influenza, and the mounting evidence that there is generalized immune-driven competition between them, it may be beneficial to simultaneously model and forecast the diseases together rather than independently [31]. Multi-disease forecasting could be achieved in future iterations of the model using similar ideas to handling multiple locations [77,78].

Overall we have shown that our model provides a disease-agnostic way to achieve robust forecast performance across seasonal diseases with a range of disease dynamics without necessitating bespoke modifications based on the disease or context. Due to the simplicity of the model and implementation, it provides a nearly off-the-shelf framework that can be rapidly deployed in new geographies and for additional seasonal diseases as desired. Such approaches will become increasingly necessary as infectious disease forecasting continues to emerge as a critical public health tool that can guide the timing and location of interventions during an outbreak [17,80].

## Methods

### Hospital admission data

We obtained data for both influenza and COVID-19 from the CDC ’s National Healthcare Safety Network (NHSN), an electronic surveillance system estimated to capture all admitted hospital patients for these diseases, though data may be backfilled over time [81]. We aggregated the daily pediatric and adult hospital admission counts at the weekly time scale for all 50 states, the District of Columbia,and Puerto Rico following the FluSight challenge methodology [18]. For influenza, we removed data prior to September 4, 2021 as there was limited influenza spread during the 2020-2021 season due to COVID-19 precautions [18,82]. For COVID-19, we used all data after October 24, 2020, as hospital reporting was sporadic before this date. We obtained RSV hospital admission counts from the Respiratory Syncytial Virus Hospitalization Surveillance Network (RSV-NET). RSV-NET is a sentinel surveillance program that includes 58 counties across 12 states (California, Colorado, Connecticut, Georgia, Maryland, Michigan, Minnesota, New Mexico, New York, Oregon, Tennessee, and Utah). RSV-NET is reported as weekly rates per 100k of the estimated population covered by each state’s RSV-NET reporting network. We therefore transformed RSV weekly rates to counts using the estimated state population coverage provided by RSV-NET. We used data after October 6, 2018 for training the model and removed data between March 1, 2020 and June 1, 2021 to remove the abnormal patterns during the initial COVID-19 transmission and resulting policies. RSV-NET only provided data for epidemiological week 40 to 18 for years prior to 2021, and we treated the summer data as missing for those years.

For the real-time FluSight challenge we made forecasts from October 14, 2023 until May 04, 2024. For our retrospective analysis for influenza and RSV we made forecasts each week from September 09, 2022 until April 21, 2023 and September 8, 2023 to April 27, 2024 to remove the summer time period. For our retrospective COVID-19 analysis we made forecasts each week from September 09, 2022 until April 27, 2024.

### The INFLAenza model

Consider a dataset of *n* observations across *M* locations and *T* timepoints. Let *y_it_* be the observed count for location *i* = 1,2,…, *M* and week *t* = 1,2,…, *T*. We model *y_it_* as a product of the expected counts per location *E_i_* and relative risk *r_it_*, where *E_i_* is a known quantity representing the number of persons at risk and *r_it_* represents latent changes over space and time. Here we use a common assumption that *E_i_* is the population size of *i*, i.e. the total state population for COVID-19 and influenza and the estimated population served for RSV, so that *r_it_* amounts to the expected risk of an individual in location *i* to be hospitalized for the disease at time *t*. Under that scenario, we model observed counts using Poisson regression as

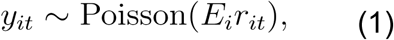

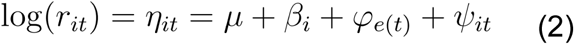

where *μ* is a global intercept, *β_i_* is a location-specific intercept, *φ_e_*_(*t*)_ is a cyclical temporal effect capturing the seasonal trend at epiweek *e*(*t*) = *t* mod 52, and *ψ_it_* is a non-cyclical spatiotemporal effect capturing short-term residual variability across both locations and time. Note that rather than assuming a specific functional form for these seasonal and short-term effects, we specify each as separate latent Gaussian processes. For the seasonal effect *φ* = (*φ*_1_,…,*φ_52_*)’, we model variability between epiweeks using a circular random walk of order two (RW2). We chose an RW2 process rather than an order 1 (RW1) model to favor a smoother curve and avoid overfitting the seasonal trend, while the circular aspect enforces that week 1 and week 52 to connect under the estimated RW2 relationship [83]. For COVID-19, which contained a training year that had 53 epidemiological weeks, we set the length of the seasonal effect to 53 instead of 52.

For the short-term residuals, *ψ*, we use a separable spatiotemporal approach [84].

Namely, each term is written as the sum of a temporal main effect and spatiotemporal interaction term, *ψ_it_* = *α_t_* + *δ_it_*. The main effect captures residual non-seasonal patterns common to all states, while the interaction term captures correlations in the deviation from *α* between states. For example, if state *i* reports an unexpected jump in hospitalizations during week *t*, we might expect a neighboring state *j* to also report a jump in cases during week *t*, or perhaps during recent or forthcoming weeks (*e.g. t* - 1 or *t* + 1). In learning these relationships from the training data, *δ* impacts the shape of forecasts for each state based on the dynamics of its neighbors.

The temporal component of both the main and interaction effects for *ψ* are modeled as a stationary autoregressive process of order one (AR1), introducing two autoregressive hyperparameters ( *ρ_α_* and *ρ_δ_*) that serve to pull predicted future values of *ψ* towards 0 in expectation (Table S2). Thus, the forecast average produced by the model will begin to revert to the seasonal trend as the length of the forecast horizon increases and short-term trends become less relevant, though *ψ* continues to impact forecast uncertainty. Finally, for the spatial part of the interaction component, we use a proper version of the Besag model, which models nearby states as more correlated than those further apart in the US neighborhood graph [85].

The shortest-path distance between pairs of states was computed using the spdep R package, which connects states based on shared borders [86]. As RSV hospital admissions are available for only 12 disconnected states, we chose to use an exchangeable spatial process, which assumes all states are equally correlated *a priori*. To fit our model and make forecasts, we use a Bayesian hierarchical framework based on Gaussian Markov Random Fields (GMRFs) made available in the R package, R-INLA [87,88].

### Prior settings and model calibration

For the global mean *μ*, we assign a highly diffuse *N(0, 30)* prior while for each location-specific intercept *β_i_* we use a standard normal distribution to avoid confounding with the global mean. Our model additionally contains 6 total hyperparameters which drive the seasonal and short-term predictions. We assign weakly-informative priors to each (Table S2). The variance parameters of the short-term main and interaction effects ( *σ_α_*^2^ and *σ_δ_*^2^) control the variability of the autoregressive process and are the most influential for calibrating forecast uncertainty. One challenge with variance parameters of hierarchical effects is that appropriate weekly-informative prior distributions depend on the scale of the data [83,89]. To address this challenge we used *penalized complexity* (PC) priors implemented in R-INLA which provide a convenient and intuitive way to adjust the priors for the variance parameters [90]. We therefore began with the default prior that is recommended for Poisson regression within R-INLA and we assessed their suitability through monitoring inference quality in two ways: (1) we inspected posterior shrinkage for the variance parameters as the difference between their prior and posterior distributions (Figure S19A), and (2) we tracked the prediction interval coverage of forecasts against their nominal expectations. Using this method, we determined that the recommended priors were appropriate for influenza and COVID-19, but that the prior scale needed to be reduced for RSV due to overall lower prevalence rates (Figure S19B).

### Model inference and forecasts

To produce forecasts for future weeks and for each state we sample from the joint posterior predictive distribution using R-INLA’s inla.jmarginal function [91]. In our case, 4-week ahead forecasts across *M* locations depend on a random variable of dimension 4 × *M*.

The joint posterior predictive distribution can also be used to produce aggregated forecasts that are ensured to be consistent across all levels of aggregation [50]. Here we produce national forecasts for influenza and COVID-19 hospital admissions using an unweighted sum over all 52 states and territories for each forecast horizon, consistent with how the national target data are produced for the FluSight challenge [92]. For RSV, we instead follow the process used by RSV-NET to produce national-level forecasts. Since the catchment area for RSV-NET is estimated to represent 9% of the national population [10], we first sampled from our model to obtain predicted counts among the population served for each of the 12 states, and then extrapolated to the national level using an unweighted sum over theses states, multiplied by 11.1 (*i.e*. 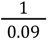).

### Forecast evaluation

We report the quality of probabilistic forecasts using two commonly used forecast metrics, the Weighted Interval Score (WIS) and the Prediction Interval Coverage (PIC) [18,21]. Since both metrics score performance based on a univariate predictive distribution and data point, we generally summarize performance as an average over a range of forecasts for different locations, dates, and forecast horizons. Both metrics are calculated with the scoringutils R package [93].

For a sequence of confidence levels *α*_1_,…,*α_K_* and predictive distribution *F*, let *l_k_* and *u_k_* denote the respective lower and upper values of a (1 - *α_k_*) x 100% prediction interval for *F*, and *m* the predicted median. The WIS is then defined for an observed true data point *y* as

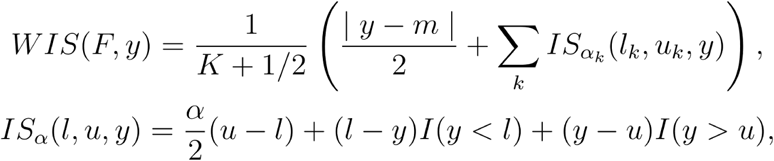

where *I* is the indicator function that takes on a value of 1 when the statement is true and 0 otherwise. WIS may be seen as a sum tallying the penalties for dispersion, overprediction, and underprediction over a larger number of prediction intervals, with lower values indicating better performance [43]. The PIC is defined in terms of a specific (1 - *α_k_*) x 100% prediction interval and confidence level *α*, and is simply equal to 1 if the observed point falls within the interval and 0 otherwise. Thus, for a sequence of forecasts and observed values across locations and horizons, the average PIC is the fraction of true values that fall within the predicted interval. A successful forecast model will therefore have a (1 - *α_k_*) x 100% PIC close to 1 - *α*. We consider 50% and 95% PICs throughout.

### Baseline reference model

We use a naive baseline model to produce forecasts that serve as a reference for the INFLAenza results, because absolute WIS values have little meaning outside of the specific forecast dataset. We use the same baseline model that is used in the COVID-19 and FluSight forecast hubs, available in the R package epipredict [18]. The model is analogous to a random walk of order 1 (RW1) with gaussian observations, where negative forecasts are truncated to zero. We calculate rWIS as the ratio between the mean WIS for the model of interest and mean WIS of the baseline. Values of rWIS below 1 indicate improvement over the baseline model. For the FluSight challenge, rWIS is calculated using a relative skill correction which adjusts the WIS of each model based on their pairwise performance between all other models, prior to normalizing WIS by the baseline model [18]. This correction is to account for the fact that not all teams submit forecasts for all locations and dates throughout the season, a problem not encountered in our retrospective analysis.

### Code

All data and code necessary to replicate our results can be found on GitHub at https://github.com/brendandaisy/inla-forecasting-paper/tree/tripledemic-paper.

## Data Availability

All code and data to reproduce the analysis are publicly available at https://github.com/brendandaisy/inla-forecasting-paper/tree/tripledemic-paper

## Acknowledgements

The authors acknowledge the helpful comments from Sam Abbott and the members of the CSTE, CDC, and MIDAS forecasting working groups. BKMC, MVS, and SJF were supported by the Council for State and Territorial Epidemiologists (NU38OT000297) and the Centers for Disease Control and Prevention (23NU38FT000008). The content is solely the responsibility of the authors and does not necessarily represent the official views of CSTE or the CDC.

