## Supplemental Information for "An accurate hierarchical model to forecast diverse seasonal infectious diseases"


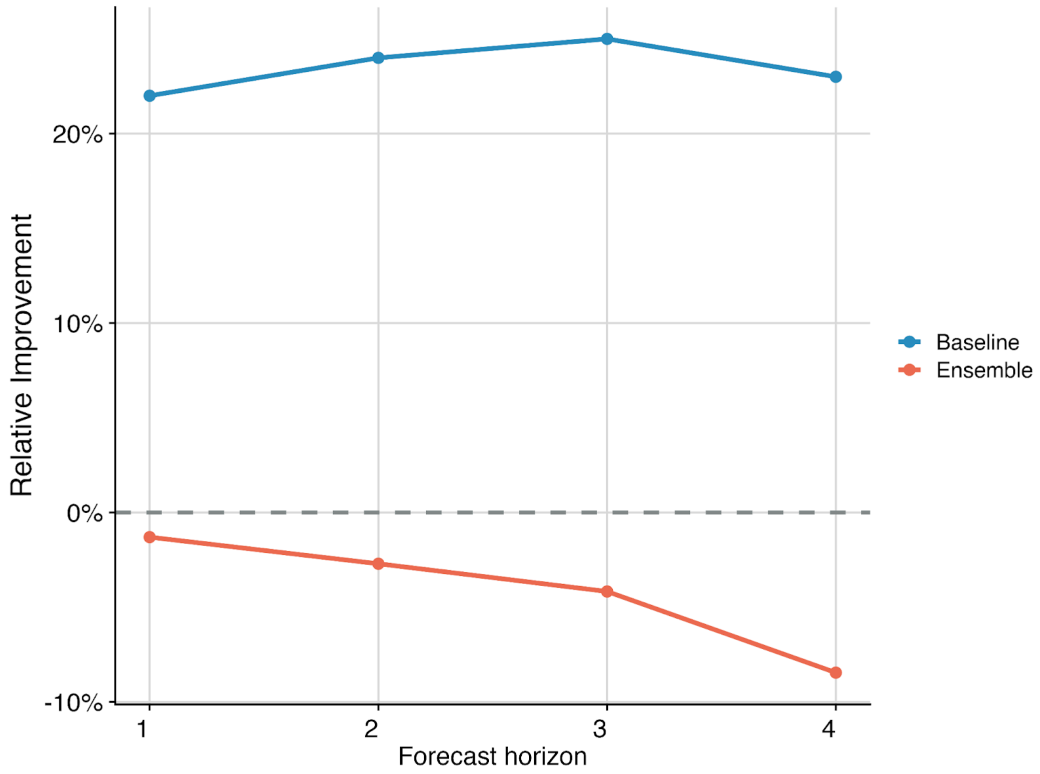


**Figure S1: Relative forecast improvement of INFLAenza by the forecast horizon compared to the Baseline and Ensemble models for the 2023-2024 FluSight challenge**. Percent improvement of the INFLAenza in WIS compared to the Baseline model (blue line and points) and Ensemble model (orange line and points). Horizontal dashed line indicates Y=0, with values above the line indicating that INFLAenza outperformed the comparative model.

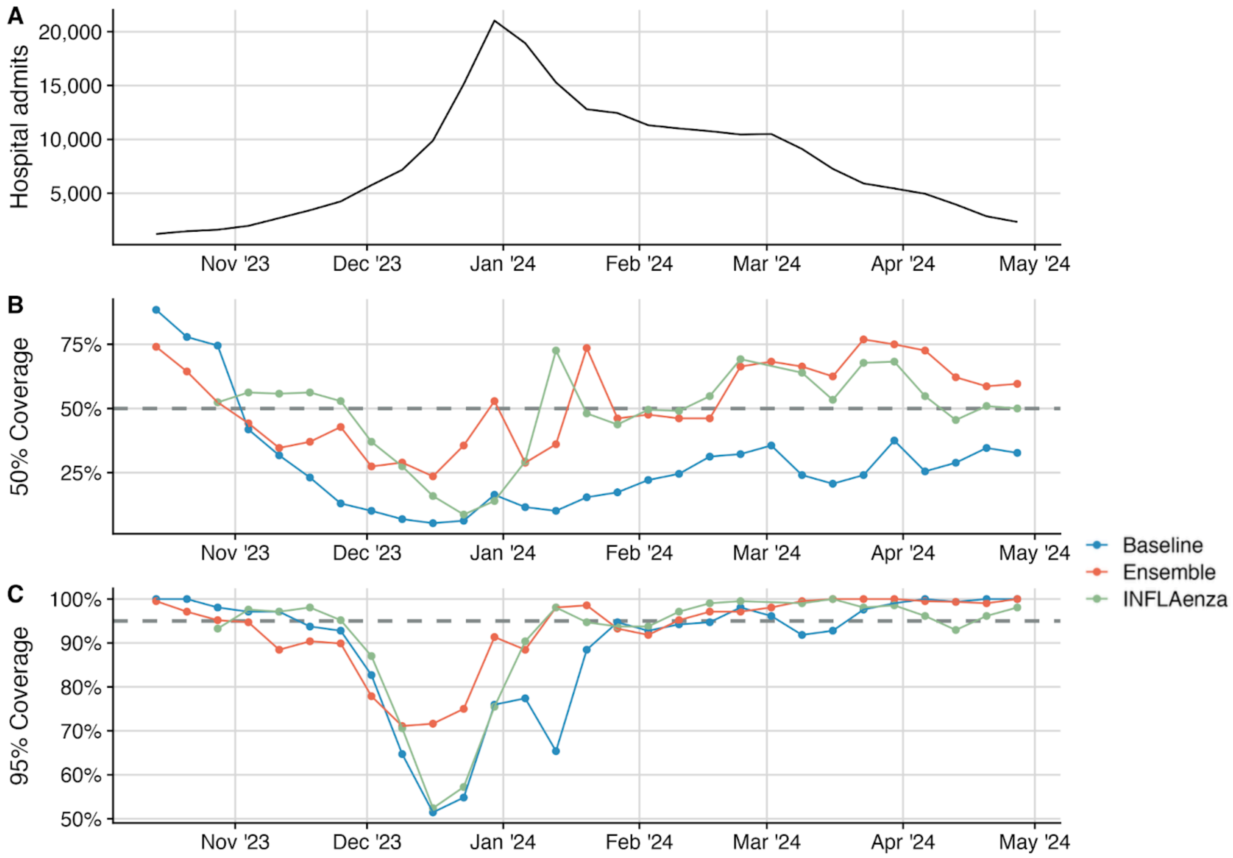


**Figure S2: Prediction interval coverage (PIC) estimates by forecast date during the 2023-2024 FluSight challenge. (A)** Weekly observed national influenza hospital admission counts from October 14, 2023, to May 04, 2024. **(B)** Comparison between the 50% PIC for the Baseline, Ensemble, and INFLAenza models (colored lines and points) against the nominal 50% coverage expectation (grey dashed line). **(C)** Comparison between the 95% PIC for the Baseline, Ensemble, and INFLAenza models (colored lines and points) against the nominal 95% coverage expectation (grey dashed line). Well calibrated models have PIC values near their nominal expectations.

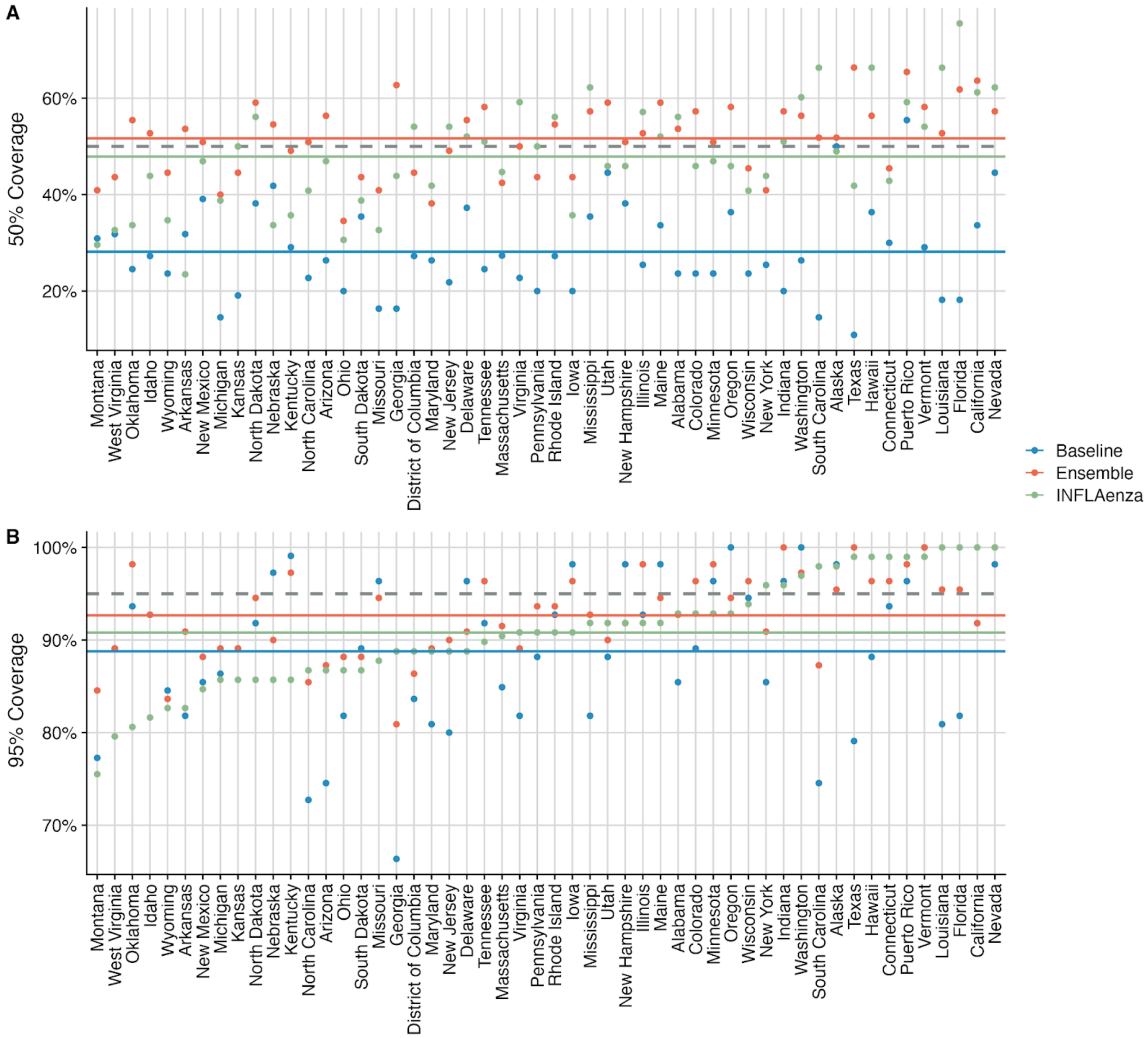


**Figure S3: Prediction interval coverage (PIC) estimates by region during the 2023-2024 FluSight challenge. (A)** Comparison between the 50% PIC for the Baseline, Ensemble, and INFLAenza models (colored points) against the nominal 50% coverage expectation (horizontal grey dashed line) for each region alongside the average PIC for each model (horizontal colored lines) **(B)** Comparison between the 95% PIC for the Baseline, Ensemble, and INFLAenza models (colored points) against the nominal 95% coverage expectation (horizontal grey dashed line) for each region alongside the average PIC for each model (horizontal colored lines). Well calibrated models have PIC values near their nominal expectations.

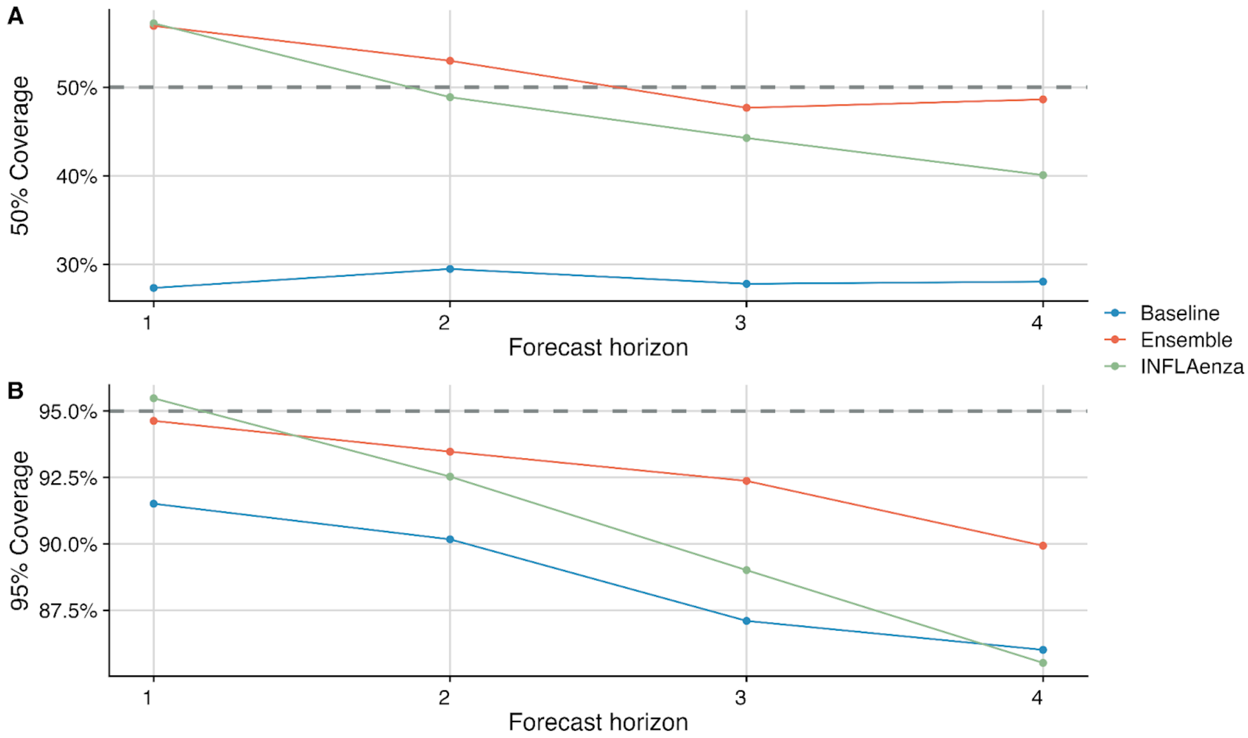


**Figure S4: Prediction interval coverage (PIC) estimates by forecast horizon during the 2023-2024 FluSight challenge. (A)** Comparison between the 50% PIC for the Baseline, Ensemble, and INFLAenza models (colored points) against the nominal 50% coverage expectation (horizontal grey dashed line) for each forecast horizon. **(B)** Comparison between the 95% PIC for the Baseline, Ensemble, and INFLAenza models (colored points) against the nominal 95% coverage expectation (horizontal grey dashed line) for each forecast horizon. Well calibrated models have PIC values near their nominal expectations.

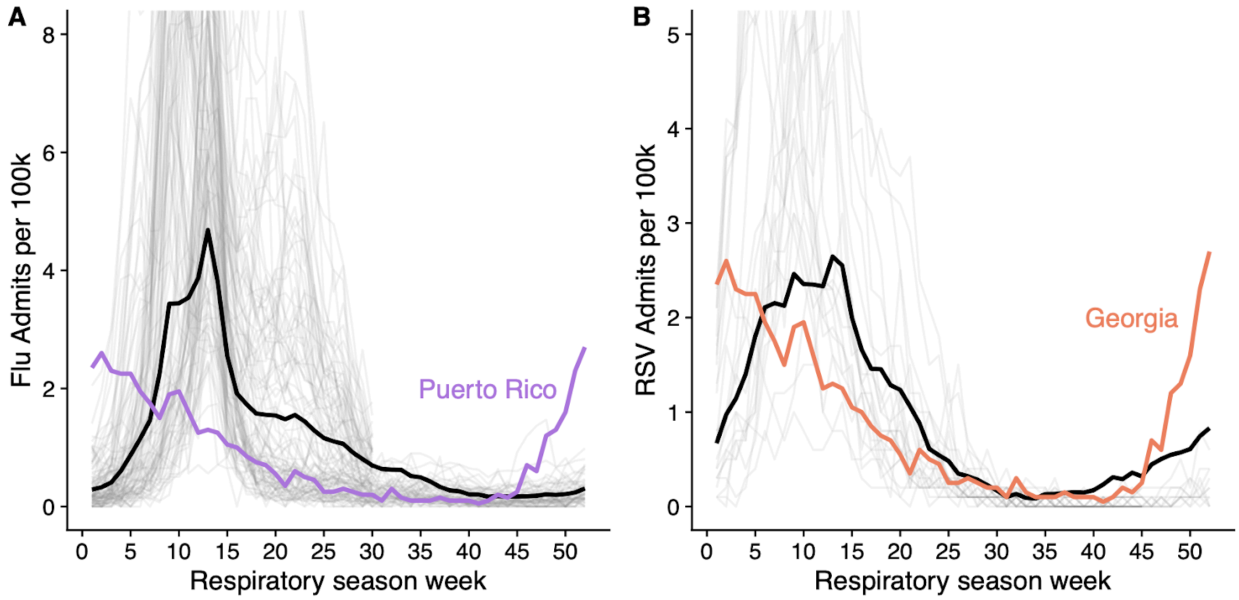


**Figure S5: Comparing seasonal patterns between the worst-performing states and the national trend for influenza and RSV.** Hospitalization rates per 100,000 individuals for each state, season, and respiratory virus (grey lines) by the week of the respiratory virus season where zero corresponds to epidemiological week 40. Black line is the weekly national average over all states and training data, and colored lines are the average admission rates of the worst performing states during the evaluated forecasting seasons (2022/23 and 2023/24).

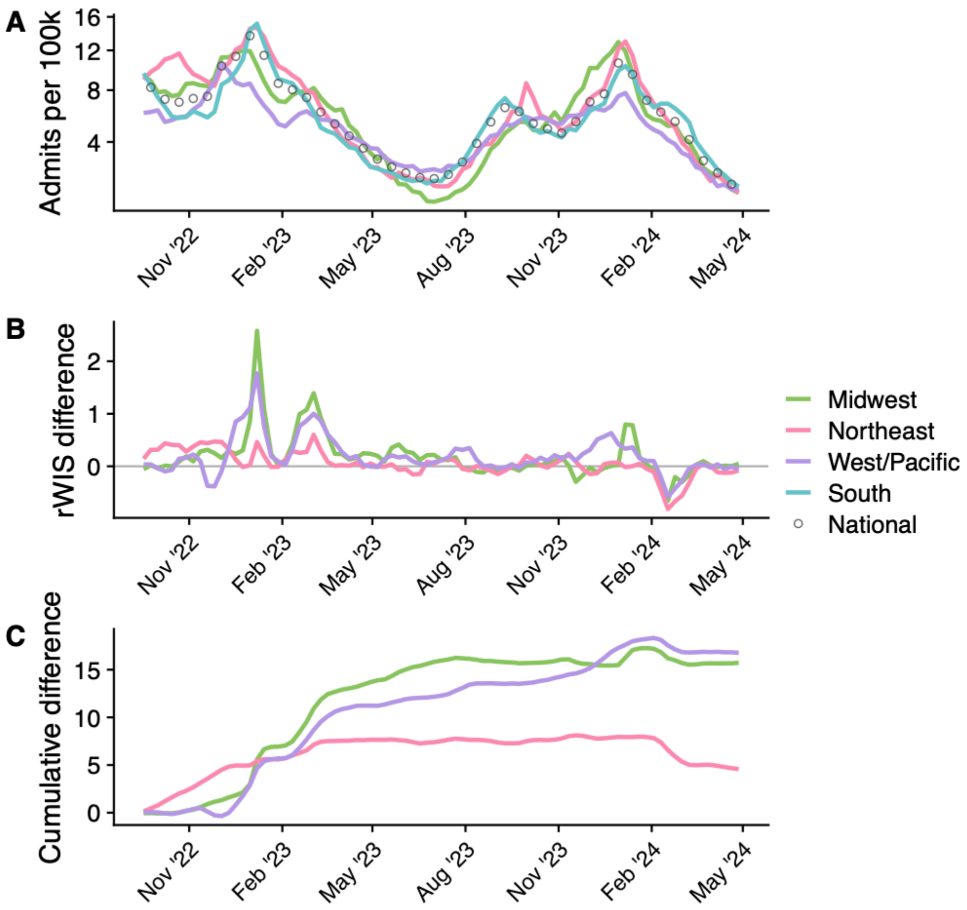


**Figure S6: Diagnosing regional forecast performance differences for retrospective COVID-19 forecasts. (A)** Confirmed COVID-19 hospital admission rates per 100,000 individuals in each US Census region (colored lines) and nationally (points), during the 2022/23 through 2023/24 respiratory seasons. **(B)** Difference between the average rWIS in the South and the three other US Census regions by forecast date (colored lines). Values above zero (horizontal grey line) indicate that forecasts for the specified region and date were worse compared to forecasts in the South relative to the baseline model. **(C)** Cumulative difference in rWIS between the specified region (colored lines) and the South over time.

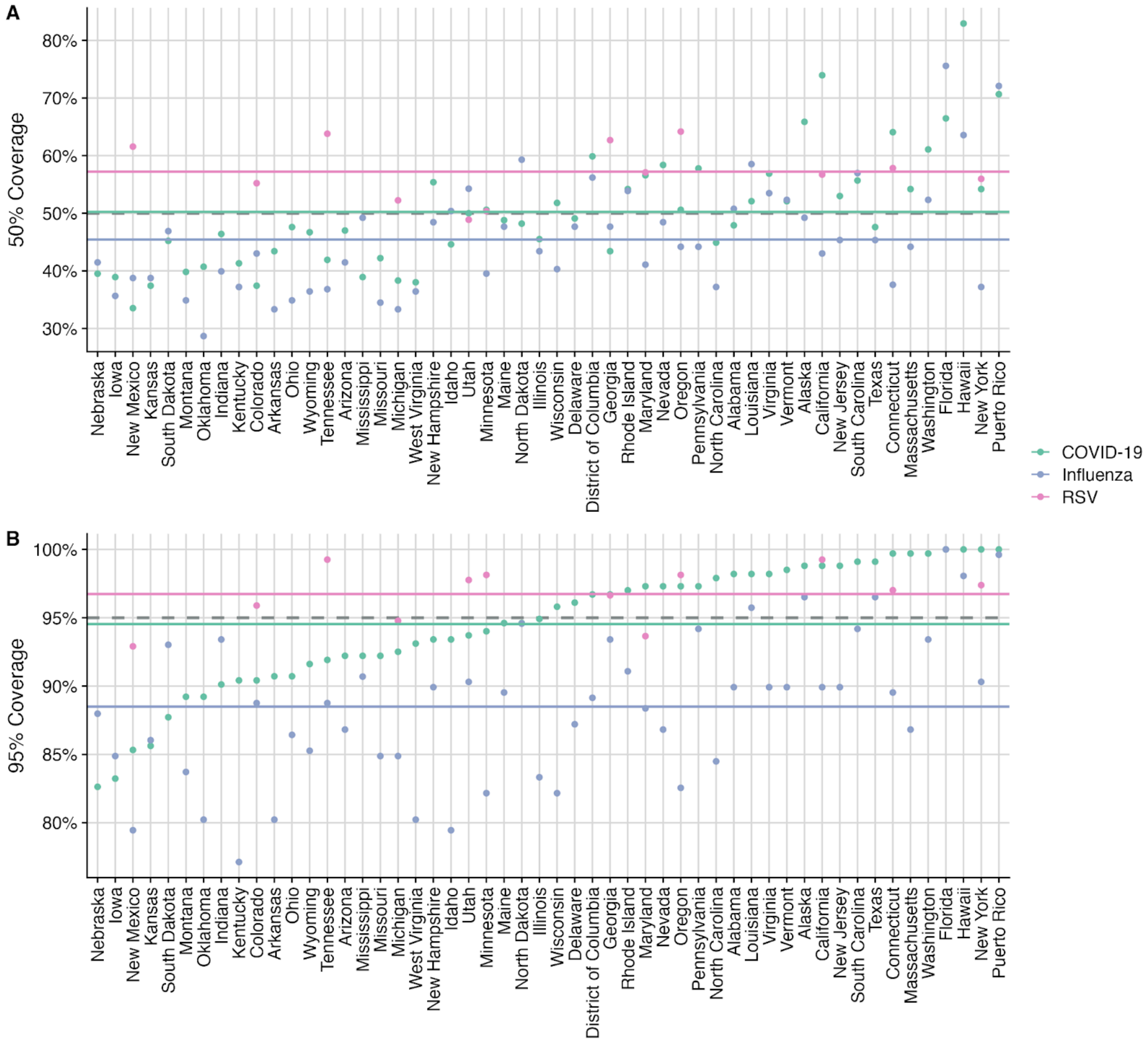


**Figure S7: Prediction interval coverage (PIC) estimates by region for retrospective forecasts of COVID-19, influenza, and RSV from September 09, 2022 to April 27, 2024. (A)** Comparison between the 50% PIC for the INFLAenza model forecasts for COVID-19, influenza, and RSV (colored points) against the nominal 50% coverage expectation (horizontal grey dashed line) for each region alongside the average PIC for each disease (horizontal colored lines) **(B)** Comparison between the 95% PIC for the INFLAenza model forecasts for COVID-19, influenza, and RSV (colored points) against the nominal 95% coverage expectation (horizontal grey dashed line) for each region alongside the average PIC for each disease (horizontal colored lines)

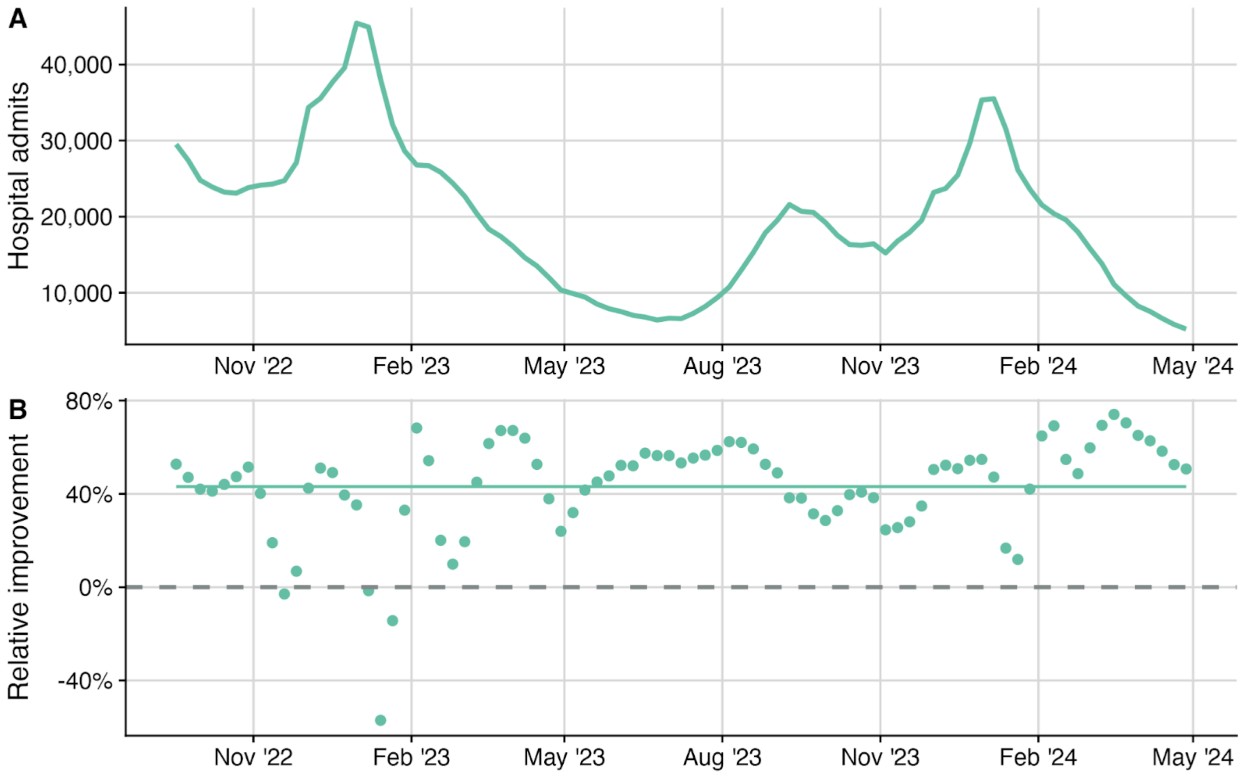


**Figure S8: Relative COVID-19 forecast improvement of INFLAenza by the forecast horizon compared to the Baseline model from September 09, 2022 to April 27, 2024**. **(A)** Weekly national COVID-19 hospital admission counts. **(B)** Percent improvement of the INFLAenza in WIS compared to the Baseline model by the date of the forecast. Horizontal dashed line indicates Y=0, with values above the line indicating that INFLAenza outperformed the Baseline model.

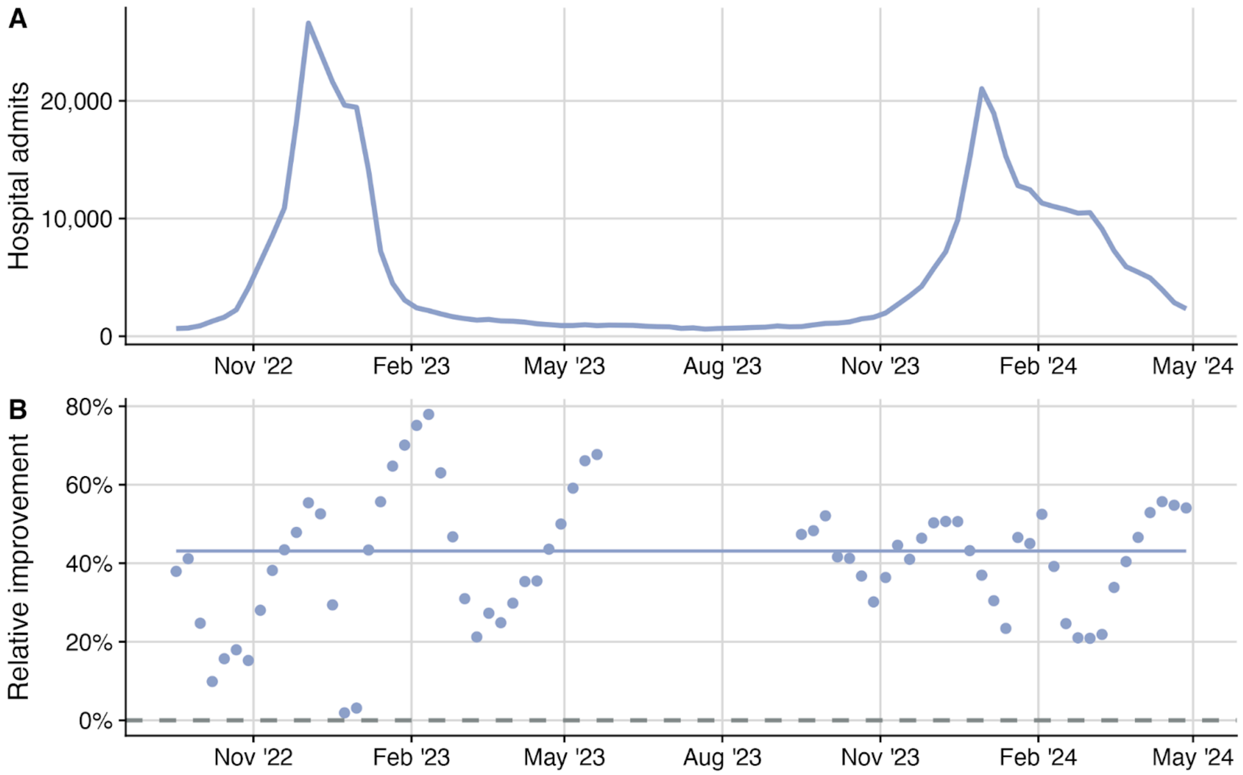


**Figure S9: Relative influenza forecast improvement of INFLAenza by the forecast horizon compared to the Baseline model from September 09, 2022 to April 27, 2024**. **(A)** Weekly national influenza hospital admission counts. **(B)** Percent improvement of the INFLAenza in WIS compared to the Baseline model by the date of the forecast. Horizontal dashed line indicates Y=0, with values above the line indicating that INFLAenza outperformed the Baseline model.

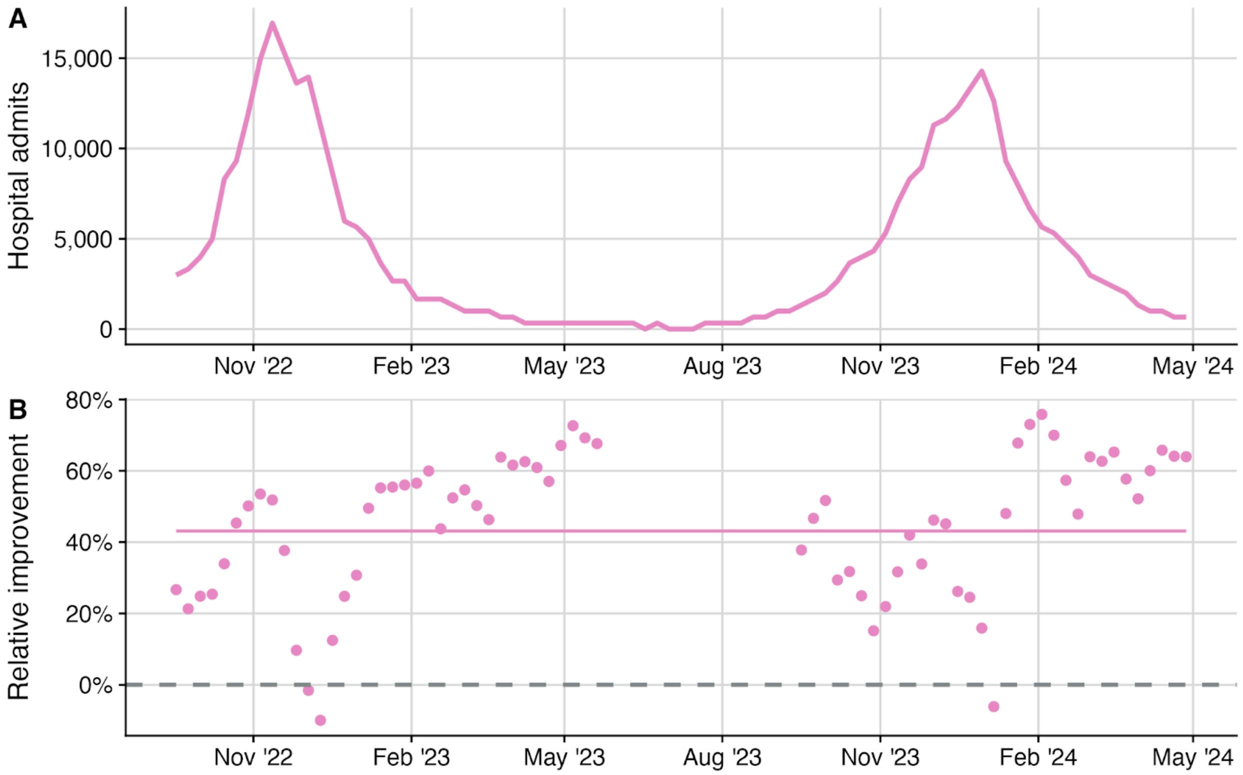


**Figure S10: Relative RSV forecast improvement of INFLAenza by the forecast horizon compared to the Baseline model from September 09, 2022 to April 27, 2024**. **(A)** Weekly national RSV hospital admission counts. **(B)** Percent improvement of the INFLAenza in WIS compared to the Baseline model by the date of the forecast. Horizontal dashed line indicates Y=0, with values above the line indicating that INFLAenza outperformed the Baseline model.

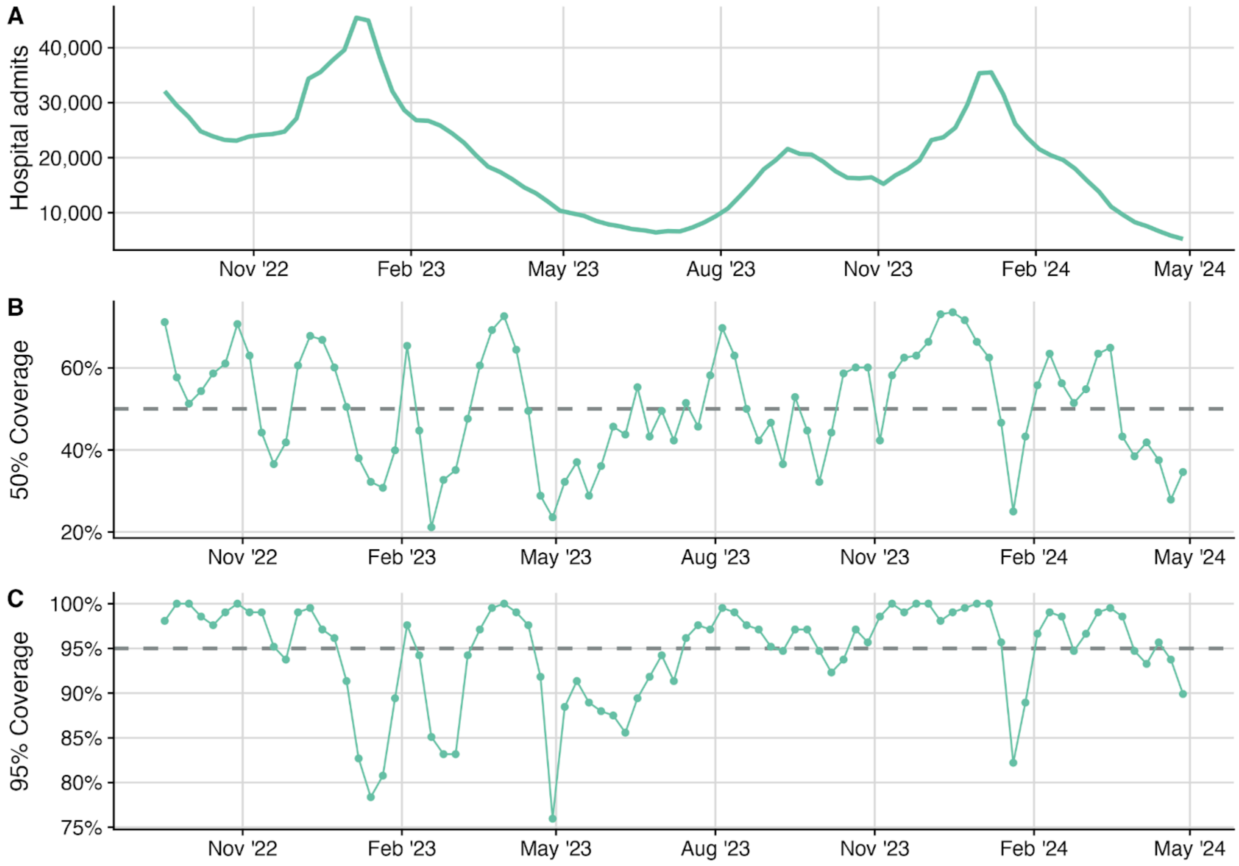


**Figure S11: COVID-19 prediction interval coverage (PIC) estimates by forecast date from September 09 2022 to April 27 2024**. **(A)** Weekly observed national influenza hospital admission counts from September 2022 to May 2024. **(B)** The 50% PIC for INFLAenza (colored line and points) against the nominal 50% coverage expectation (grey dashed line). **(C)** The 95% PIC for INFLAenza (colored line and points) against the nominal 95% coverage expectation (grey dashed line). Well calibrated models have PIC values near their nominal expectations.

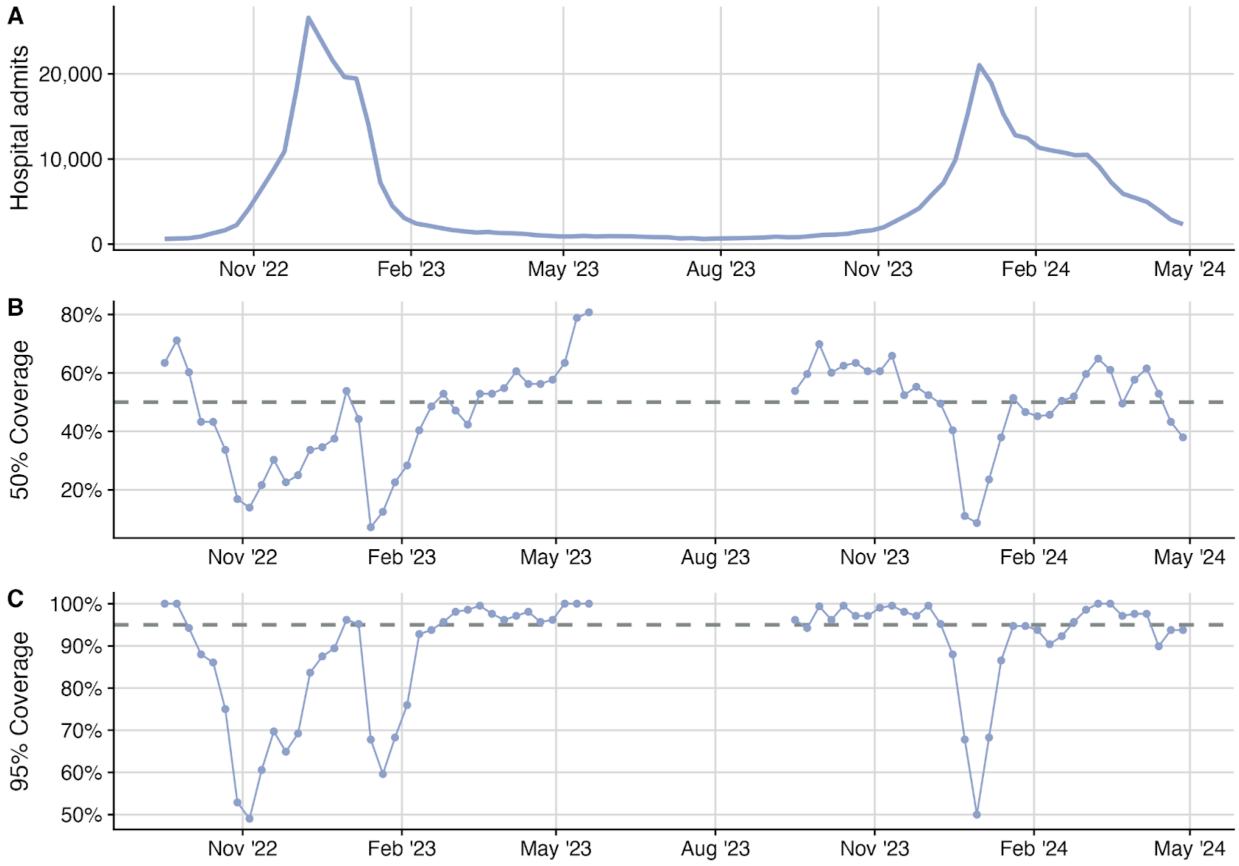


**Figure S12: Influenza prediction interval coverage (PIC) estimates by forecast date from September 09 2022 to April 27, 2024**. **(A)** Weekly observed national influenza hospital admission counts from September 2022 to April 2024. **(B)** The 50% PIC for INFLAenza (colored line and points) against the nominal 50% coverage expectation (grey dashed line). **(C)** The 95% PIC for INFLAenza (colored line and points) against the nominal 95% coverage expectation (grey dashed line). Well calibrated models have PIC values near their nominal expectations.

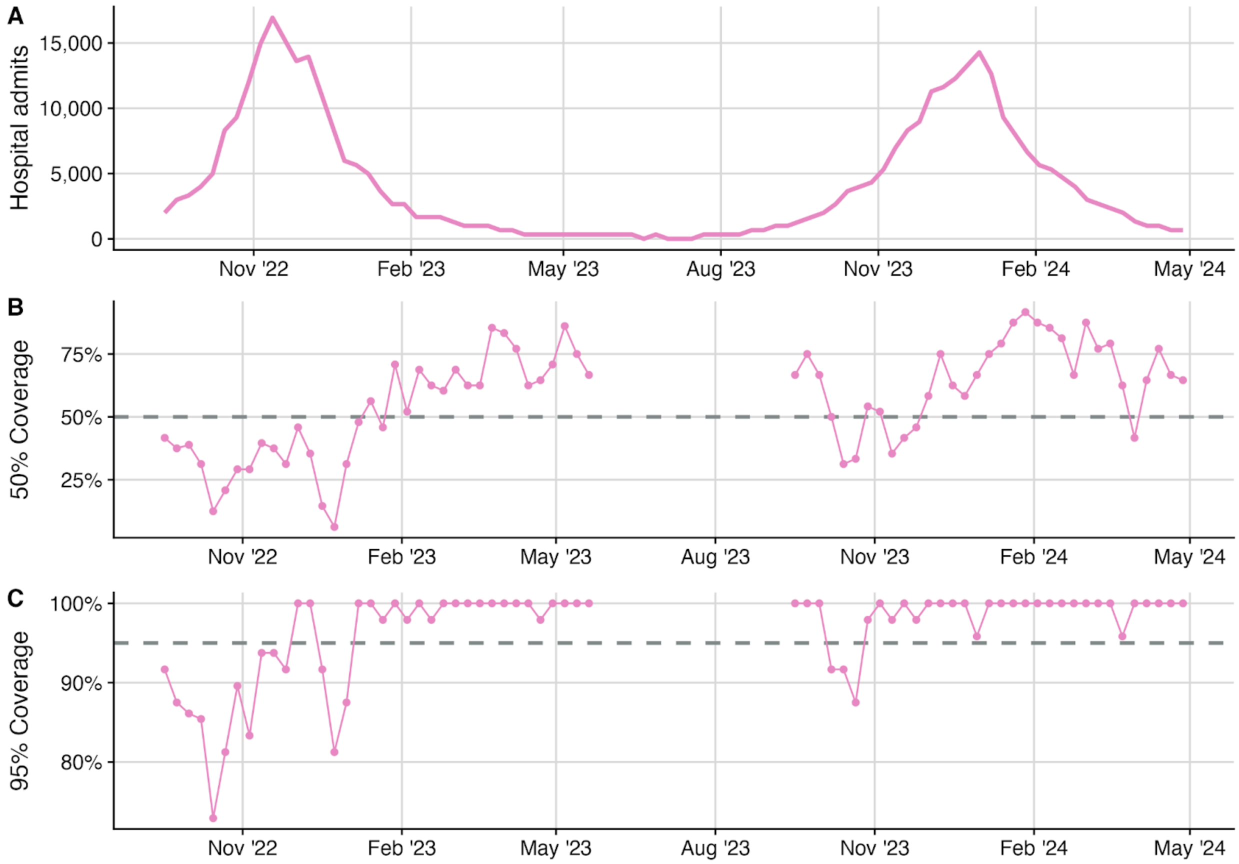


**Figure S13: RSV prediction interval coverage (PIC) estimates by forecast date from September 09, 2022 to April 27, 2024**. **(A)** Weekly observed national influenza hospital admission counts from September 2022 to April 2024. **(B)** The 50% PIC for INFLAenza (colored line and points) against the nominal 50% coverage expectation (grey dashed line). **(C)** The 95% PIC for INFLAenza (colored line and points) against the nominal 95% coverage expectation (grey dashed line). Well calibrated models have PIC values near their nominal expectations.

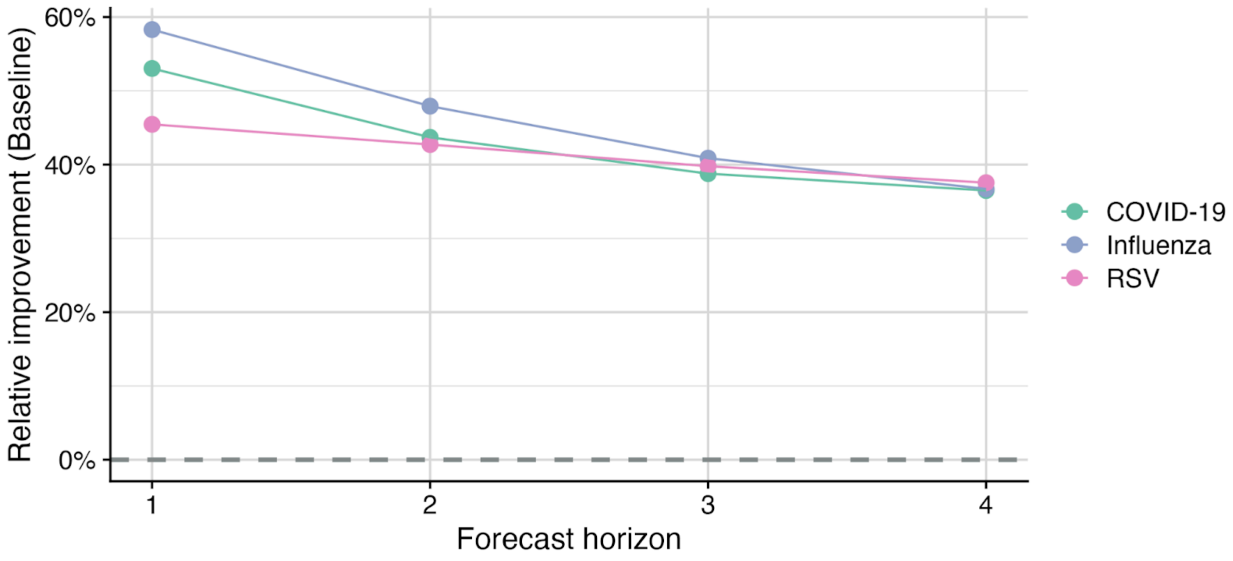


**Figure S14: Relative forecast improvement of INFLAenza by the forecast horizon compared to the Baseline model from September 09, 2022 to April 27, 2024**. Percent improvement of the INFLAenza in WIS compared to the Baseline model for each disease (colored lines and points). Horizontal dashed line indicates Y=0, with values above the line indicating that INFLAenza outperformed the Baseline model.

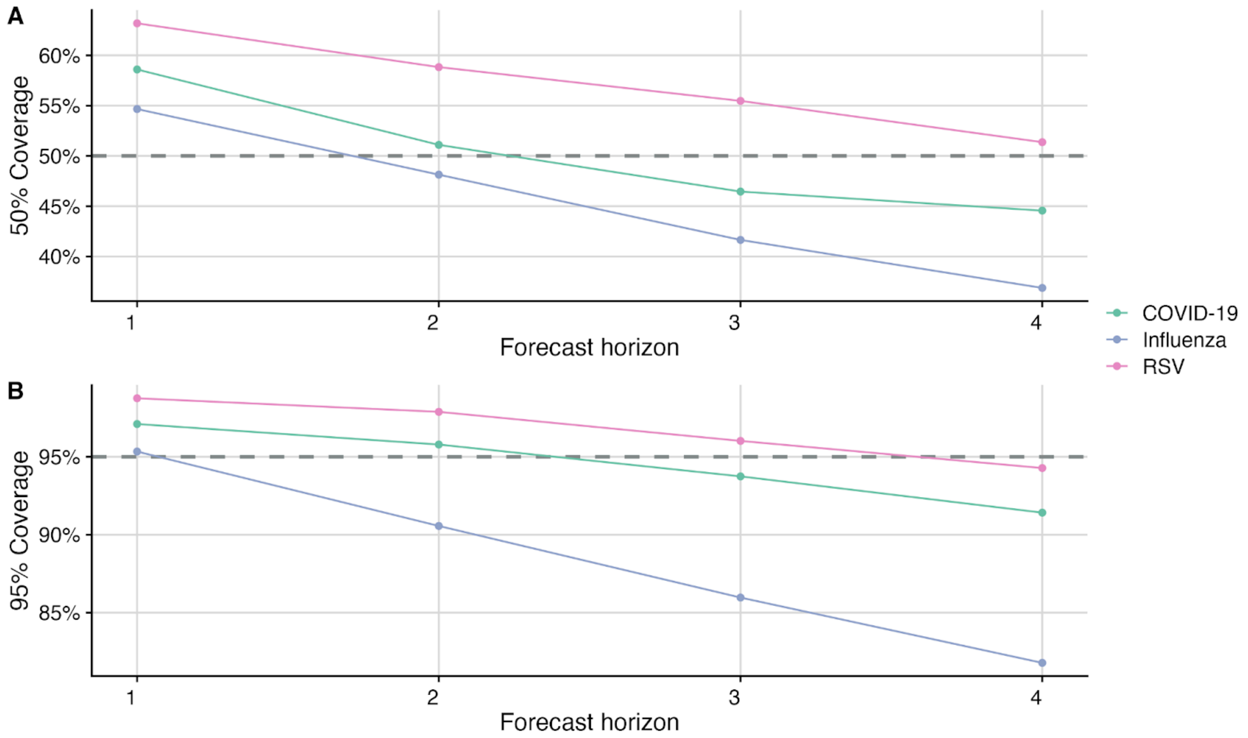


**Figure S15: Prediction interval coverage (PIC) estimates by forecast horizon for COVID-19, influenza, and RSV forecasts. (A)** Comparison between the 50% PIC for the COVID-19, influenza, and RSV forecasts (colored points and lines) against the nominal 50% coverage expectation (horizontal grey dashed line) for each forecast horizon. **(B)** Comparison between the 95% PIC for the COVID-19, influenza, and RSV forecasts (colored points and lines) against the nominal 95% coverage expectation (horizontal grey dashed line) for each forecast horizon. Well calibrated models have PIC values near their nominal expectations.

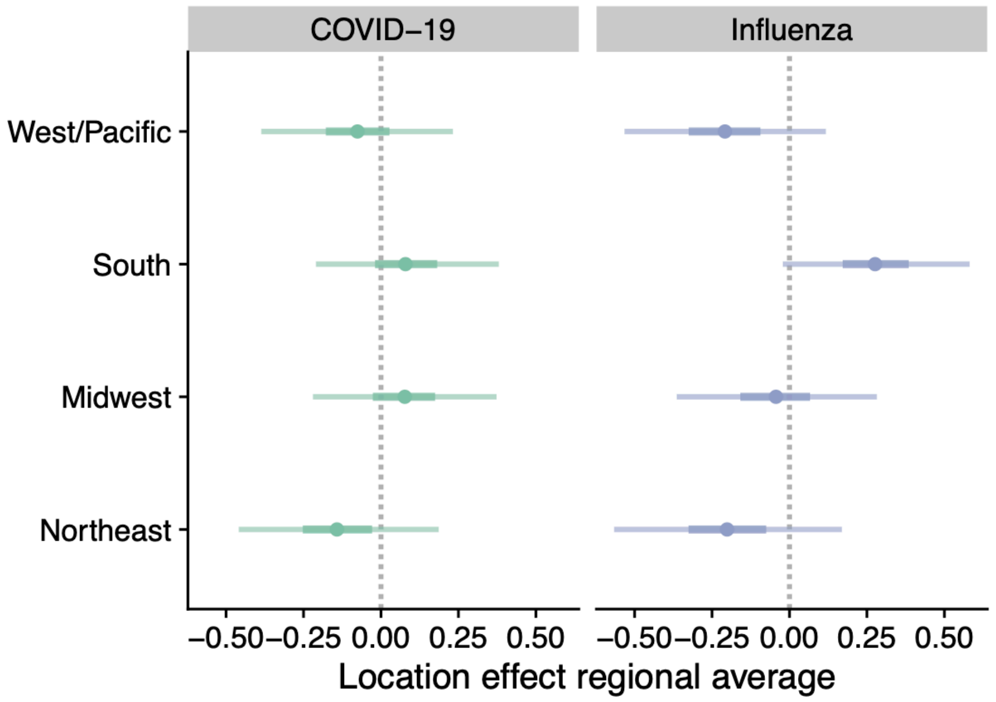


**Figure S16: Estimated COVID-19 and influenza hospital admission burden summarized by United States census region.** Distributions for the regional averages are derived using posterior estimates of the location-specific intercepts ($\beta$ in Eq. (2)) by drawing 5,000 samples from the joint distribution over $\beta$, then taking the average by region for each sample. The resulting distributions are displayed as points (median), thick lines (75% CrI), and thin lines (95% CrI). Posterior estimates are derived using all training data up until April 27, 2024. Regions are defined using the 4 US Census statistical regions, with Puerto Rico and the District of Columbia added to the South region. Values below zero indicate an overall aggregate lower rate of hospital admissions compared to the national average throughout the time series.

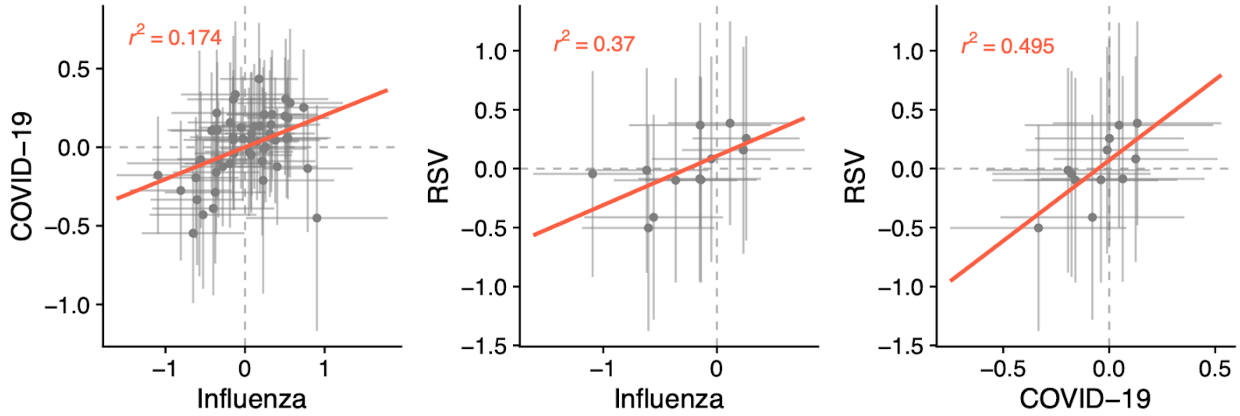


**Figure S17: Scatterplots between the inferred location-specific hospital admission burden for comparisons between the COVID-19, influenza, and RSV models.** Posterior mean for the location-specific intercepts (points, $\beta$ in Eq. (2)) and 95% credible intervals (horizontal/vertical lines), with best-fit regression line between the diseases (red line). $r^2$ is the coefficient of determination. Posterior estimates are derived using all training data up until April 27, 2024. Values below zero (vertical and horizontal dashed lines) indicate an overall lower rate of hospital admissions compared to the national average throughout the time series for the specific disease and location.

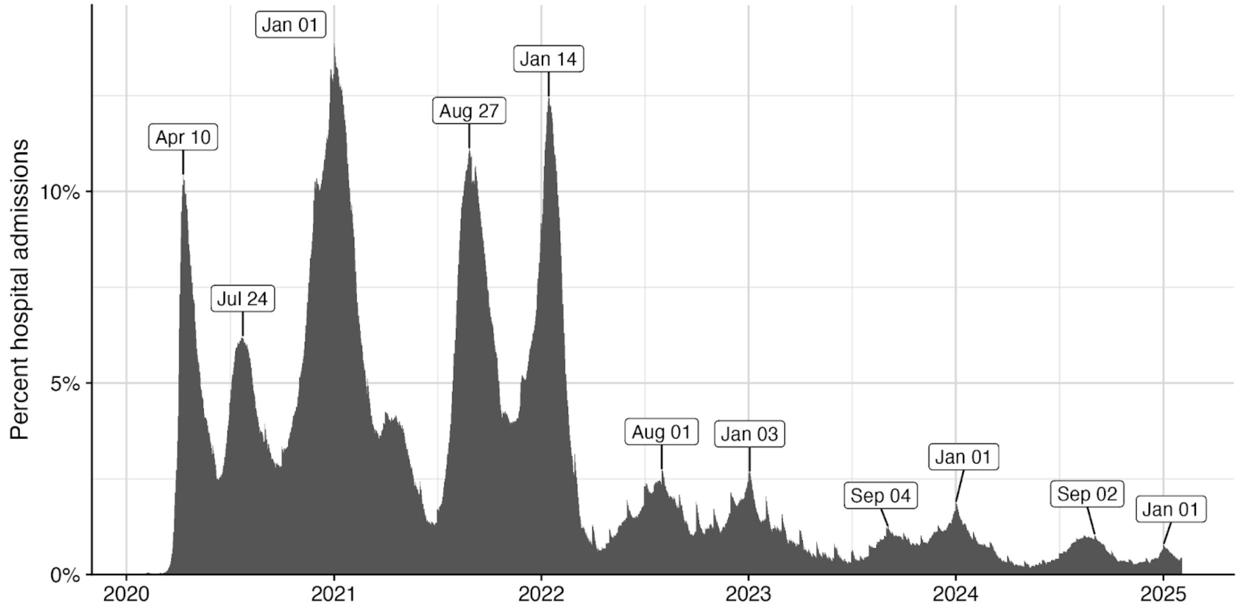


**Figure S18: Percentage of daily hospital admissions attributed to COVID-19 from electronic health records claims data from January of 2020 until February of 2025.** Labels identify the timing of the maximal daily value for each major peak during the time period, highlighting the biannual COVID-19 epidemic over five years.

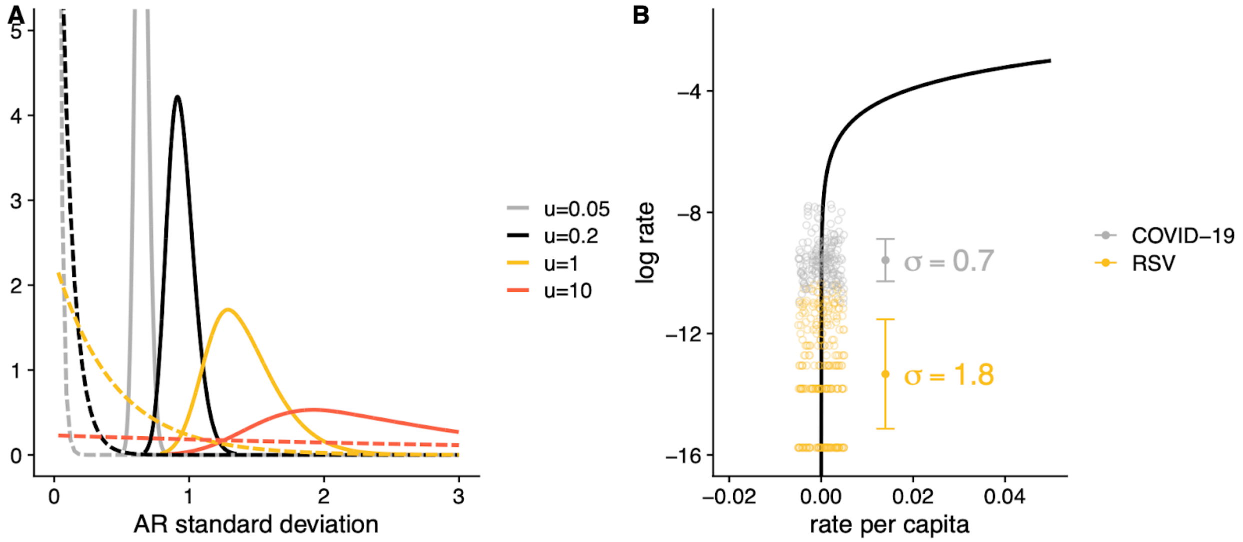


**Figure S19: Adjusting the precision of temporal effects to the scale of the data for RSV.** **(A)** The prior (dashed) and posterior (solid) distributions of the marginal standard deviation of an AR1 process fit to the RSV data. Prior distributions are $PC(u, 0.05)$ for varying values of $u$, where PC is the penalized complexity prior of Simpson et al. (2017). Starting with the most diffuse prior ($u$=10), we find natural support within the data for a standard deviation above 1, but that the posterior upper tail is poorly constrained by the data. In this case we identify ($u$=0.2) as an appropriate choice as it is the most informative prior that maintains probability mass near the lower bound posterior estimate from the most diffuse prior. **(B)** Hospitalization rates for RSV and COVID-19 data mapped to log scale. Error bars indicate mean $\pm$, where $\sigma$ is the empirical standard deviation. The prevalence of very low rates in the RSV data can be seen to overly inflate the empirical standard deviation, as these values approach $-\infty$ on log scale.

| Disease | Model Variation | WIS | rWIS | 50% Coverage (%) | 95% Coverage (%) |
|---|---|---|---|---|---|
| COVID-19 | **Full model** | 48.7 | 0.64 | 50.1 | 94.4 |
| | **No seasonal** | 57.4 | 0.75 | 46.0 | 94.0 |
| | **No spatial** | 59.8 | 0.78 | 50.3 | 94.2 |
| Influenza | **Full model** | 40.7 | 0.61 | 45.4 | 88.3 |
| | **No seasonal** | 44.8 | 0.68 | 46.7 | 85.5 |
| | **No spatial** | 42.0 | 0.63 | 47.4 | 87.5 |
| RSV | **Full model** | 9.0 | 0.65 | 56.3 | 96.3 |
| | **No seasonal** | 9.8 | 0.71 | 50.4 | 93.8 |
| | **No spatial** | 9.2 | 0.67 | 49.9 | 93.0 |

**Table S1: Retrospective performance of the full INFLAenza model compared to simpler variants.** 4-week ahead forecasts were produced weekly for each model and disease, and dates were the same as the retrospective analysis in the main text, ranging from September 9, 2022 to April 27, 2024. The "no seasonal" model was the same as the proposed model, but with the seasonal effect ($\phi$ in Eq. (2)) removed, while the "no spatial" model removed the location intercepts and spatiotemporal interaction term ($\boldsymbol{\beta}$ and $\delta$) while fitting the remaining parameters independently for each state. In the main text, percentage improvement of simpler model variants compared to the full model is computed as the average relative improvement over the three diseases.

| Symbol | Description | Prior |
|---|---|---|
| $\tau_\varphi$ | precision of the seasonal RW2 effect | $\tau_\phi \sim PC(1, 0.05)$ |
| $\rho_\alpha$ | autoregressive parameter of the short-term main effect | $\log \frac{1+\rho_\alpha}{1-\rho_\alpha} \sim N(0, 6.67)$ |
| $\tau_\alpha$ | marginal precision of the short-term main effect | $\tau_\alpha \sim PC(1, 0.05)$ |
| $\rho_\delta$ | autoregressive parameter of the short-term interaction effect | $\log \frac{1+\rho_\delta}{1-\rho_\delta} \sim N(0, 6.67)$ |
| $\tau_\delta$ | marginal precision of the short-term interaction effect | $\tau_\delta \sim PC(1, 0.05)$ |
| $d$ | degree of properness/inverse strength of neighborhood correlations | $\log d \sim \text{Gamma}(1, 1)$ |
| $r$ | correlation between each location's timeseries (exchangeable version) | $\log \frac{1+r(M-1)}{1-r} \sim N(0, 5)$ |

Table S2: Hyperparameters and their default priors in the model. Note priors for several parameters are specified on a transformed scale. $PC$ = penalized complexity prior of Simpson et al. (2017). $M$ = number of locations.
